# The Mathematics of Serocatalytic Models with applications to public health data

**DOI:** 10.1101/2025.01.21.25320864

**Authors:** Everlyn Kamau, Junjie Chen, Sumali Bajaj, Nicolás Torres, Richard Creswell, Jaime A. Pavlich-Mariscal, Christl Donnelly, Zulma Cucunubá, Ben Lambert

## Abstract

Serocatalytic models are powerful tools which can be used to infer historical infection patterns from age-structured serological surveys. These surveys are especially useful when disease surveillance is limited and have an important role to play in providing a ground truth gauge of infection burden. In this tutorial, we consider a wide range of serocatalytic models to generate epidemiological insights. With mathematical analysis, we explore the properties and intuition behind these models and include applications to real data for a range of pathogens and epidemiological scenarios. We also include practical steps and code in R and Stan for interested learners to build experience with this modelling framework. Our work highlights the usefulness of serocatalytic models and shows that accounting for the epidemiological context is crucial when using these models to understand infectious disease epidemiology.

## 1 The information contained in age-structured serological data

In a serological survey, specimens from a sample population are tested for antibodies for a pathogen. Although direct results from these tests provide a continuous measurement of the level of antibodies in an individual, results are typically categorised into binary outcomes: an individual is *seropositive* if their antibody response is above a threshold and *seronegative* otherwise ^1^.

Serological surveys (also known as *serosurveys*) are typically cross-sectional, taken at a point in time. From these surveys we can calculate the proportion of individuals who are seropositive, which we term *seroprevalence* or alternatively *seropositivity*, and we use each of these terms interchangeably throughout this paper.

In Figure 1, we show data from three yellow fever serosurveys in the Americas presented in Hugo Muench’s classic 1934 study ^2^, work which popularised serocatalytic modelling. Whereas the two Brazilian datasets generally display an upward trend in seroprevalence with age, the Colombian seroprevalence is generally flat across ages.

**Figure 1:**
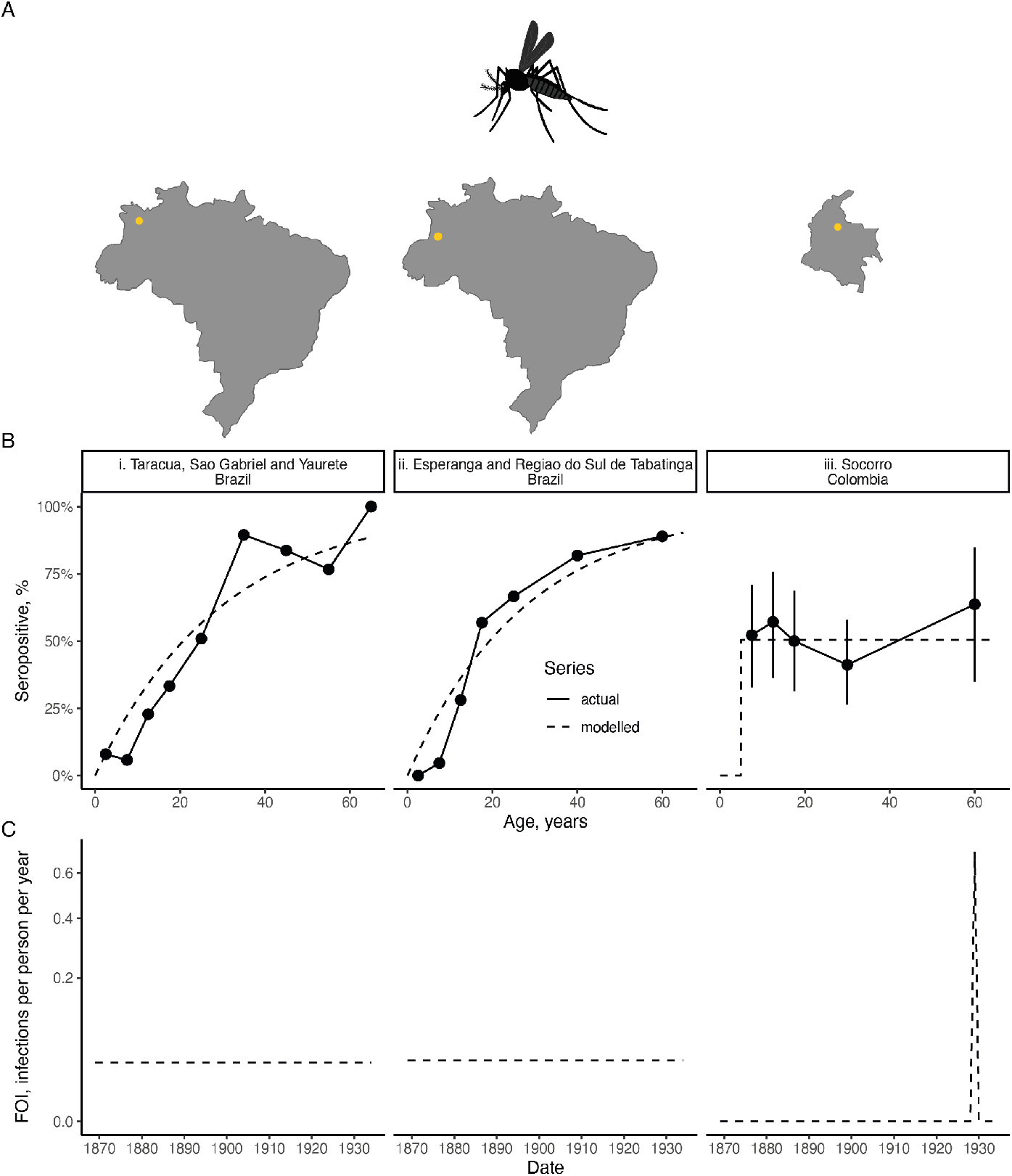
Muench’s serosurvey data for yellow fever from three locations in the Americas. Row A shows the locations of the serosurveys. Each column in rows B and C represents a particular location in the Americas. Row B shows the raw data (black points connected by solid lines) and modelled proportions seropositive (dashed lines); in column iii, we show the 2·5th and 97·5th percentiles of a posterior distribution assuming a uniform prior over the proportion seropositive and a binomial likelihood for each age group separately; in columns i× ii, we do not show uncertainty intervals since we did not have access to the sample sizes used in the serosurveys. Row C shows the inferred historical FOIs. Details of the data and analysis are provided in §A.1.

Faced with these patterns, Muench asks the question: what transmission patterns generated these data? He argues that the two datasets from Amazonian Brazil (Figure 1B, i & ii) were generated from a transmission history distinct from that of Colombia. Running contrary to popular opinion at the time, his claim is that yellow fever was effectively *endemic* in these locations in Brazil. In Sorocco, Colombia, however, it was known that the town had, in living memory, been free from yellow fever until an isolated outbreak occurred in 1929; that is, it was known that transmission of yellow fever was effectively *epidemic* in this region.

Transmission rate is quantified through a metric known as the *force of infection*, or FOI, the rate at which individuals become infected per unit time; FOI has units of the average number of infections a person receives per unit time (usually per year) – note, this metric can exceed 1 if it is expected that an individual would be infected more than once in a given time interval. Endemic transmission then translates into assuming that the FOI varies little over time. This assumption results in a characteristic increase in seroprevalence as individuals age, and in Figure 1B, i & ii, we show the estimated seroprevalence resulting from assuming an FOI that does not vary over time. The estimated equivalents (dashed lines) generally provide a reasonable approximation to the underlying trends in the raw data (solid points and lines).

For an isolated epidemic, assuming everyone sampled in the population was exposed to the same degree results in a constant proportion seropositive for those old enough to have experienced the epidemic and survived it. We show the estimated seropositive proportion under this assumption in Figure 1B (iii).

In Figure 1C, we then show the estimated FOIs. Under these assumptions, the historical FOIs in the two Brazilian locations were virtually identical, and the inferred FOI for the 1929 Colombian yellow fever outbreak is far in excess of these.

This example illustrates the power of serological data: by making assumptions about the nature of the transmission dynamics, we can reveal unobserved historical transmission patterns. These patterns can be used to project future transmission dynamics and, based on seroprevalence levels, inform vaccination strategies. Serological data can play an important role in disease surveillance, particularly in circumstances where surveillance is challenging; for example, for novel pathogens with unknown or nonspecific symptoms or in locations with limited resources.

What serological data tell us about transmission patterns hinges on assumptions about how, if at all, transmission varies over time. It also depends on assumptions about exposure patterns across different demographics, and whether individuals lose antibody detectability over time – otherwise known as *seroreversion*.

These assumptions are embodied by mathematical models referred to as *serocatalytic* models, and this article provides an introductory guide to this class of models.

## 2 Reproducing our methods and results

This paper is meant as an accessible guide for serocatalytic modelling and promotes learning through step-by-step mathematical derivations, simulation of models and fitting these to data to infer transmission dynamics. Table 1 summarises the models covered and structure of this article.

**Table 1:**
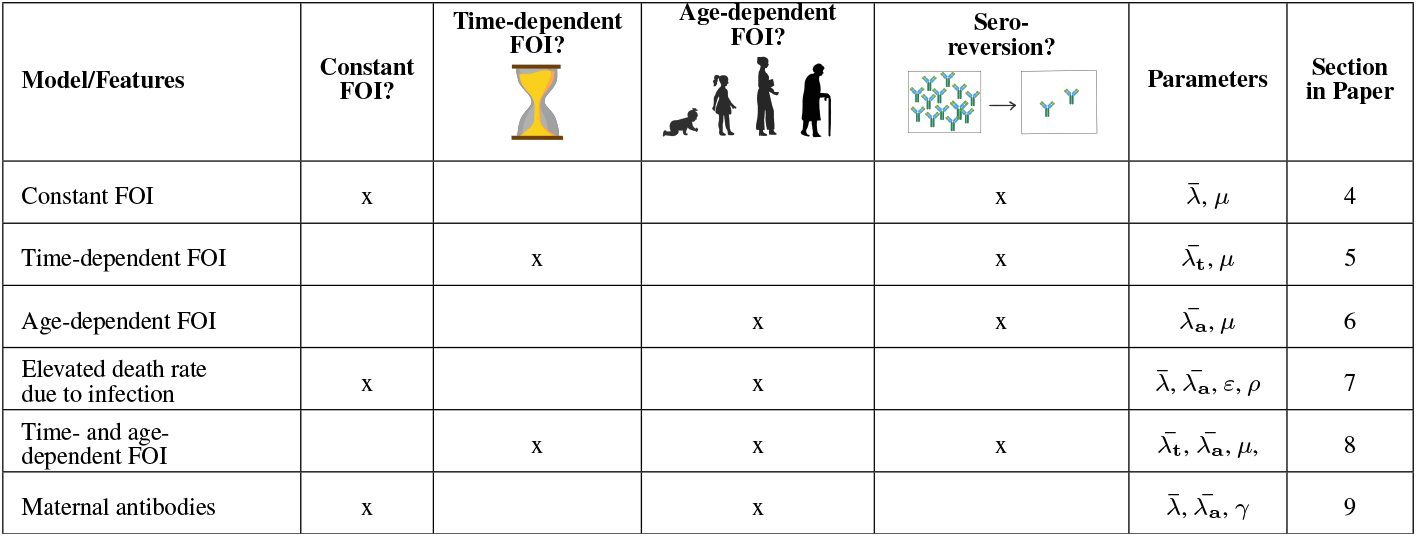
Summary of the serocatalytic models introduced in this study. The models explored here are listed in the first column, and the corresponding features available with each model derivation marked with “x” in the respective cells. The images shown in the three middle columns show the recurring pictorial representations that we use to represent these classes of model. Also shown are notations for parameters that can be inferred in each model. The section describing each model is listed in the last column. FOI means the force of infection.

| Model/Features | Constant<br>FOI? | Time-dependent<br>FOI? | Age-dependent<br>FOI? | Sero-<br>reversion? | Parameters | Section<br>in Paper |
| --- | --- | --- | --- | --- | --- | --- |
| Constant FOI | x | | | x | $\bar{\lambda}, \mu$ | 4 |
| Time-dependent FOI | | x | | x | $\bar{\lambda}_t, \mu$ | 5 |
| Age-dependent FOI | | | x | x | $\bar{\lambda}_a, \mu$ | 6 |
| Elevated death rate<br>due to infection | x | | x | | $\bar{\lambda}, \bar{\lambda}_a, \varepsilon, \rho$ | 7 |
| Time- and age-<br>dependent FOI | | x | x | x | $\bar{\lambda}_t, \bar{\lambda}_a, \mu$ | 8 |
| Maternal antibodies | x | | x | | $\bar{\lambda}, \bar{\lambda}_a, \gamma$ | 9 |

A summary of the mathematical symbols used throughout is provided in Glossary S1. A key notational rule we use throughout is to use lowercase symbols to denote continuous quantities (e.g. *b*, the specific calendar date/time at which birth occurs) and uppercase symbols to denote integer values (e.g. *B*, the year in which individuals were born).

We demonstrate the behaviour of each model by simulating from it in R ^3^. The simulated dynamics are shown in figures throughout the paper, and we provide a website which includes R code and further visualisations (https://arianajunjie.github.io/seropackage/).

We also illustrate the application of serocatalytic models by fitting actual serological data of various pathogens, with code available from https://github.com/ekamau/serocatalytic_models. For the model fitting, the parameters of each model were estimated through a Bayesian framework. More detailed information on the inference is provided in §A.3. For our model fitting, we used the *targets* ^4^ R package to create reproducible data analysis pipelines and *renv* ^5^ to allow our computational environment to be reproducible by others.

## 3 Transmission dynamics models and their relationship with serocatalytic models

Transmission dynamics models attempt to mechanistically model the population dynamics of pathogens through host populations. Many such models are built upon the classic Susceptible-Infected-Recovered (SIR) model, with the following form:

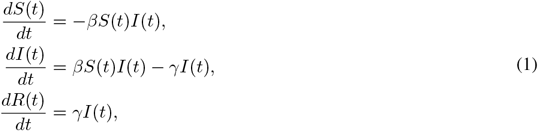

where *S*(0) = *S*_0_, *I*(0) = *I*_0_, *R*(0) = *R*_0_ are the initial conditions. We assume *S*(*t*) + *I*(*t*) + *R*(*t*) = 1, for *t*≥ 0, meaning we model the dynamics of the proportion of individuals who are susceptible, (*S*(*t*)), infected (*I*(*t*)) and recovered (meaning they cannot be reinfected; *R*(*t*)).

Models such as eq. (1) attempt to mechanistically describe how an epidemic system unfolds with time. But this comes at a cost – in order to predict future system behaviours, it is necessary to have reasonable knowledge about the system. To solve eq. (1), we need to know the transmission rate, *β >* 0, the rate of recovery from infection, *γ >* 0, and the initial conditions. For novel pathogens or during an outbreak, these parameter values may be unknown.

Further, SIR models like eq. (1) are highly idealised, and this model system does not standardly allow *β* or *γ* to vary over time; for example, in response to the imposition of public health interventions or due to the introduction of novel pathogen variants. Eq. (1) is also inherently a short-term model system since it fails to account for births or natural deaths which collectively change the susceptible population. The model also does not include seroreversion or account for loss of antibody detection, where individuals once recovered become susceptible or *seronegative*. For a review of the various ways in which simple SIR-type models may be extended to answer a variety of epidemiological questions, see ^6^.

Transmission dynamics models have been extraordinarily successful, but their correct use requires incorporating substantial epidemiological knowledge. Serocatalytic models instead limit the reach of the modelling. Whereas some classes of transmission dynamics models can be used to predict numbers of future infections, serocatalytic models cannot be used directly to predict how an epidemic might unfold into the future – they inherently look into the past.

Whereas transmission dynamics models track the status of a population through time, including the proportions infected (Figure 2A), serocatalytic models instead attempt to explain and quantify the rates at which existing individuals in a population have historically become infected throughout their lifetimes (Figure 2C). Whereas transmission dynamics models provide a mechanistic basis for *how* infected individuals contribute to ongoing transmission, serocatalytic models do not directly model the drivers of ongoing transmission but instead are useful to infer *when* infections occurred.

**Figure 2:**
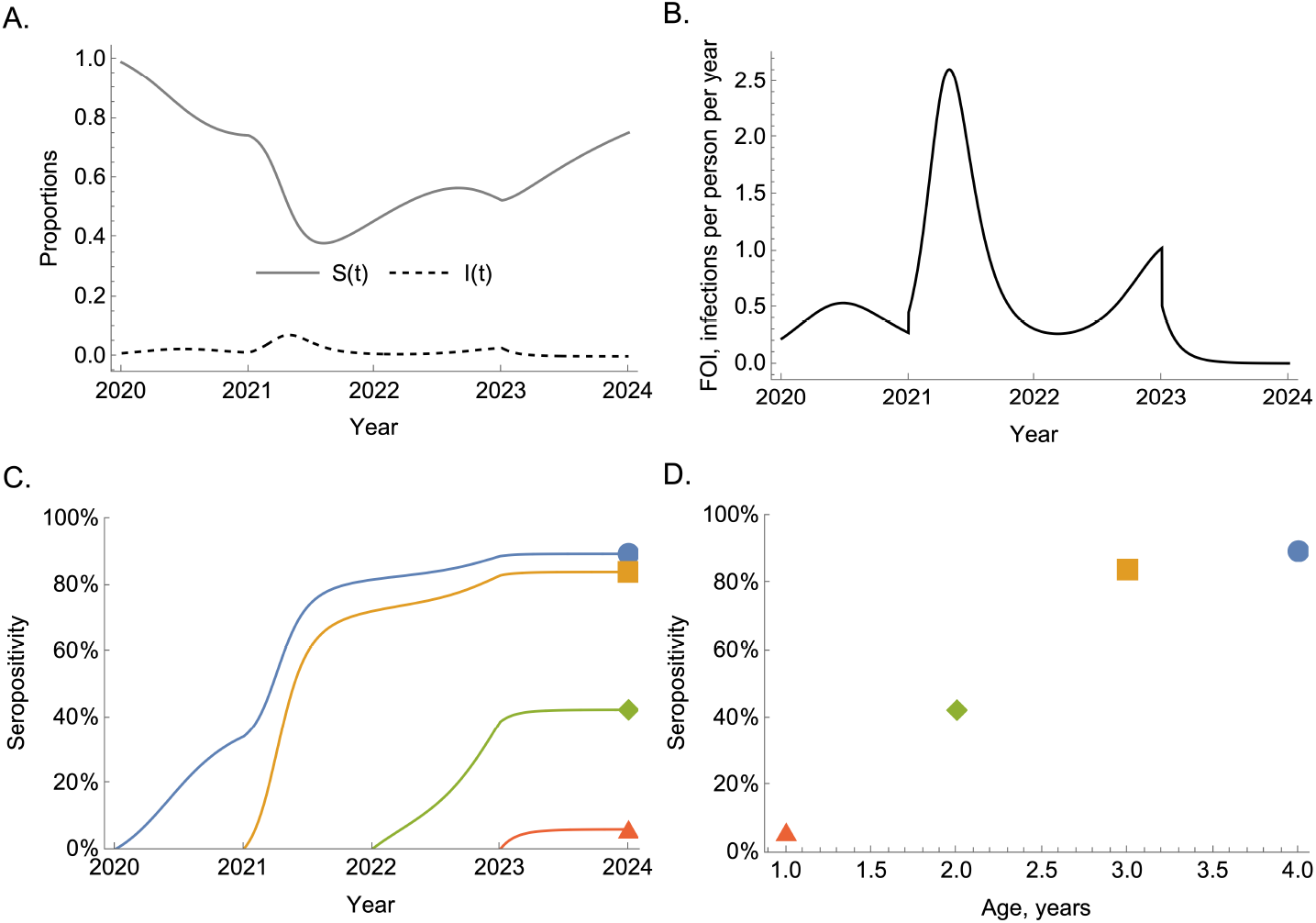
The relationship between transmission dynamics models and serocatalytic models. Panel A shows the proportions susceptible and infected resultant from simulating a transmission dynamics model that includes waning immunity. In this model, the transmission rate, *β*, varies over time (see §A.2 for a complete description). Panel B shows the force of infection: *β*(*t*)*I*(*t*). Panel C shows the seropositivity trajectories of four birth-cohorts. Panel D shows the serological age profile, in years, in the population in 2024.

We can convert the SIR example of eq. (1) into a serocatalytic model by effectively ignoring the relatively short-term dynamics of the infected population and considering infection dynamics within just two groups: susceptible (i.e. seronegative) individuals, *S*(*t*), and *seropositive* individuals, *X*(*t*) – those previously infected, which includes both currently infected and recovered individuals. To remove dependence on the numbers of infected individuals and to generalise the system to allow for time-varying transmission rates, we introduce *λ*(*t*) = *β*(*t*)*I*(*t*), where *λ*(*t*) is the FOI (Figure 2B). Eqs. (1) then simplify to:

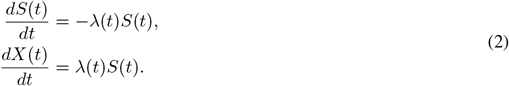

However, Eqs. (2) require us to know an initial condition: the proportion of susceptible individuals at some time in the past, represented by 0 ≤ *S*(0) ≤ 1, which may not be known if no serosurveys were conducted at that time. For this, serocatalytic models both simplify and complicate the problem simultaneously – they abandon the idea of modelling the whole population and instead consider birth-cohorts, individuals born in the same year. Whilst this means that we get one set of equations for each birth-cohort, we gain an initial condition by assuming that each birth-cohort is seronegative at birth (see ^2^). This means that, unlike transmission dynamics models, serocatalytic models are *always* age-structured. We assume that each birth-cohort has the same birth-time, *b* ≤ *t*.

For each cohort then, we have equations of the form:

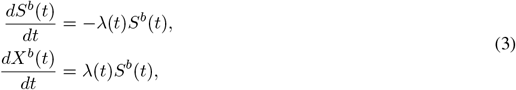

where 0 ≤ *S*^*b*^(*t*) ≤ 1 is the proportion of individuals born at time *b* who are seronegative at time *t* and have initial conditions: *S*^*b*^(*b*) = 1, *X*^*b*^(*b*) = 0 meaning that *S*^*b*^(*t*) + *X*^*b*^(*t*) = 1 for all *t* ≥ *b*.

Using *S*^*b*^(*t*) = 1 − *X*^*b*^(*t*), we can rewrite the second equation for those seropositive in Eqs. (3) as:

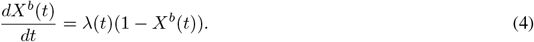

By solving eq. (4) for each birth-cohort, we can then determine their serological trajectories through time (Figure 2C); in §4 we explain how this equation can be solved. This allows us to determine a cross-sectional profile of seropositivity stratified by age at a particular point in time (Figure 2D); note that the age profile of seropositivity may vary for serosurveys conducted at different points in time.

### 3.1 Population-level versus individual-level quantities

FOI represents the average number of infections a person receives per unit time, and the typical time unit is years; it is a rate, meaning it must be zero or above and has no upper bound. An alternative way to represent this quantity is to convert it into the probability that an individual becomes infected in a particular year. To obtain this quantity we consider the risk that an individual becomes infected within a year. If the FOI within a year is constant at 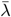,the probability an individual remains uninfected (*U*) throughout that year can be calculated by solving:

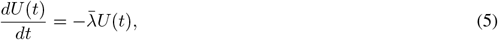

with *U* (0) = 1 meaning the individual starts in an uninfected state; there is a negative sign in eq. (5) because individuals flow from being uninfected to being infected over time. To solve this for the probability they are uninfected at the end of year, we use the method of separation of variables to yield: 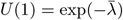 meaning the probability they are infected is 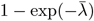.

More generally, when using serocatalytic models, there is a distinction between population quantities and individual quantities. The seroprevalence is the proportion of individuals who are seropositive in the population; the analogous quantity for a randomly chosen individual is the probability that they are seropositive, and these two quantities will be numerically equal and are given by *X*^*b*^(*t*). For example, if the seroprevalence is 50%, the probability a randomly chosen individual is seropositive is 0.5.

## 4 Constant force of infection

We first consider an idealised situation where the FOI does not vary over time, representing *endemic* transmission of the form Muench claimed could result in the transmission patterns for yellow fever in the Brazilian Amazon ^2^; see §1. This type of FOI could not be generated by a transmission dynamics model like that described by eq. (1) because it permits no non-zero disease equilibrium, but a simple modification of this equation to include births of susceptible individuals and/or waning immunity would allow this ^6^.

Assuming the FOI is fixed at 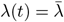,eq. (4) becomes:

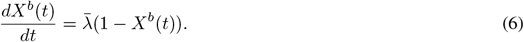

We can then separate the variables:

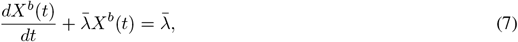

and employ the “integrating factor” approach to solve differential equations by multiplying both sides of eq. (7) by 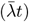 and rewriting the left-hand-side as a derivative:

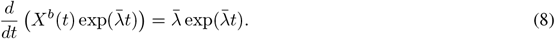

We then integrate both sides from *b*, when individuals were born, to *t > b*. To do so, we introduce a dummy integration variable *t*^′^:

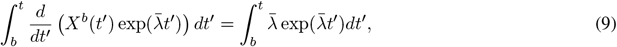

which leads to the following:

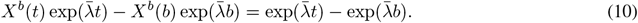

Using the initial conditions, we can rearrange eq. (10) to obtain the proportion seropositive at time *t*:

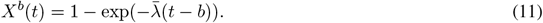

We can use eq. (11) for a range of birth-cohorts to determine their seropositivity trajectories through time (to produce seropositivity profiles like those shown in Figure 2).

### 4.1 Seropositivity by age

Another way to write eq. (11) is to introduce a birth-cohort-specific age, *a*^*b*^ := *t* − *b*, for any *t* ≥ *b*, and *b* is constant:

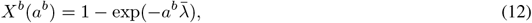

which shows that, as individuals age, their seropositivity increases due to their cumulative risk of infection.

We can also consider seropositivity across a population at a snapshot at time *t*. For this, we introduce a variable, *a*_*t*_ = *t*− *b* to denote the age of individuals at time *t*; note that, unlike for our birth-cohort-specific age, *b* varies since individuals in the population may be of different ages. We can then use eq. (12) to determine how seropositivity varies across the population:

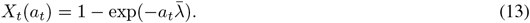

While eq. (12) and eq. (13) appear similar, they represent quite different quantities: the former gives the seropositivity trajectory for a particular birth-cohort born at time *b*, for example, like the trajectories shown in Figure 2C; the latter gives the average seropositivity for an individual of a specific age at time *t* (i.e. across all birth-cohorts born prior to that year) – see Figure 2D for an example of such a serological cross-section.

## 5 Time-varying force of infection

A more general situation is when the FOI has varied historically. It can be assumed that the FOI is a continuous function of time, but, for many practical purposes, ages of individuals in serological surveys are recorded in calendar years. So, often a sensible simplifying assumption is that the FOI is piecewise-constant, typically with pieces of one year in length. In what follows, we assume this unless explicitly stated otherwise.

Under the piecewise-constant assumption, the FOI experienced by a cohort born at the start of year *B* until year *T > B* is given by a series of yearly FOIs: 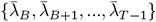.For example, *λ*_*B*_ is the FOI level experienced continuously from the start of year *B* until its end.

When an FOI is constant within a year, we can directly integrate eq. (8) between the start of a year and its end. We illustrate this between the year of birth, *B*, and the following year; in doing so, we assume individuals are born at the start of year *B*:

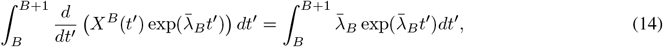

which yields the following solution for the proportion seropositive one year after their birth:

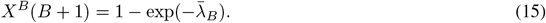

Repeating this for the next year, we have:

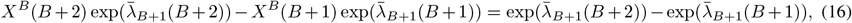

which can be rearranged to:

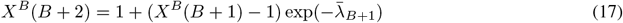

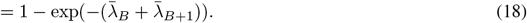

Iteratively, for an arbitrary year *T > B*, we find that:

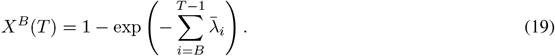

Eq. (19) effectively defines a survival model, where the probability of becoming infected by a given time relates to the cumulative force of infection to which an individual has been exposed. If there is strong transmission throughout an individual’s life, they are more likely to have been infected and be seropositive by the time of antibody measurement.

It is also possible to reparameterise eq. (19) in terms of a birth-cohort-specific age: *A*^*B*^ = *T*− *B* (in integer years), where *B* is fixed and *T* varies:

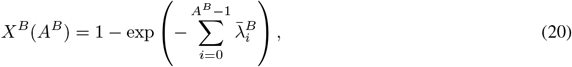

where 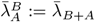 is the force of infection experienced by a cohort born in year *B* as a function of its age. When transmission varies over time, different birth-cohorts can experience varied histories of transmission. This means that each age group will experience its own set of 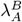 values. For example, suppose that those aged one at the start of 2024 (i.e. those born at the start of 2023) were subject to a constant force of infection 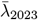 in their first year of life. This means the proportion seropositive will be:

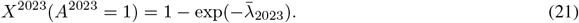

Those aged two at the start of 2024 will experience the same force of infection in their second year as those aged one did in their first, *λ*_2023_. But they will have also experienced a force of infection *λ*_2022_ in their first year of life, meaning the proportion seropositive of this cohort at the start of 2024 is given by:

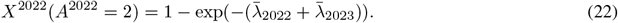

We now consider the seropositivity profile in a given year *T* as a function of integer age *A* of individuals within a population: this emulates the typical data obtained from serological surveys, which can be obtained from eq. (19):

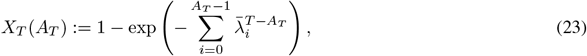

where *A*_*T*_ = *T* − *B* corresponds to the age of a cohort born at the start of year *B*. We use the notation *A*_*T*_ to denote the age of a member of the population surveyed in year *T*, i.e. *B* varies across the population; this is distinguished from *A*_*T*_ which is the age of a particular birth-cohort at time *T*, i.e. *B* is fixed.

In Figure 3, we illustrate the impact of a time-varying FOI by considering a range of birth-cohorts for a pathogen with varied historical transmission rates (panel A). Panel B shows the solution given by eq. (19) for six different age cohorts: in each case, we solve the model from their birth until the present time (taken as *T* = 2024 in eq. (19)). In panel C, we show the seropositivities of each age cohort in 2024 as calculated by eq. (23).

**Figure 3:**
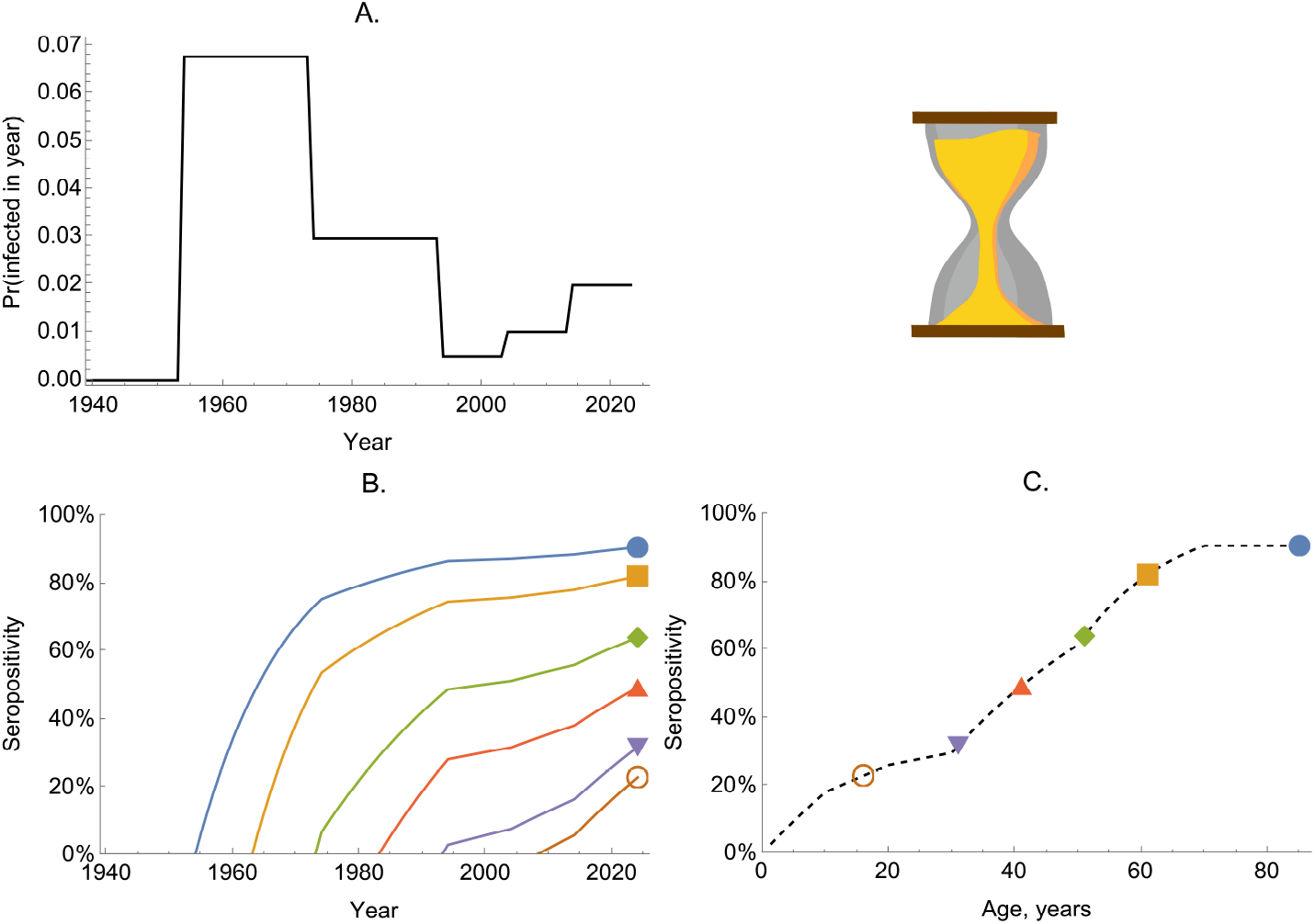
The dynamics of seropositivity in a time-varying FOI model. Panel A shows the probability of becoming infected in a given year (given by 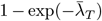) over time. Panel B shows the solution of eq. (19) for six birth-cohorts (coloured lines). Panel C shows the proportion seropositive by age for the population in 2024; the dashed line shows the solution for all age groups and the coloured markers correspond to the seropositivities shown in 2024 in panel B.

### 5.1 Including seroreversion

Until now we have assumed that antibodies, as a signature of past infection, are always detectable. For some infections, however, antibody levels wane with time below the limit of detection and *seroreversion* is said to have occurred, defined in this context as a confirmed positive serologic test later testing negative ^7,8^. We now discuss how to incorporate seroreversion into serocatalytic models by building on the time-varying model introduced above.

We modify eq. (3) to allow seropositive individuals to become seronegative:

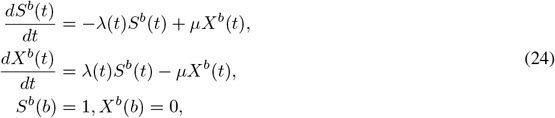

and assume that the rate of seroreversion, *µ >* 0, is constant. Since *S*^*b*^(*t*) + *X*^*b*^(*t*) = 1 for *t*≥ *b*, we can rewrite the equation for seropositive individuals:

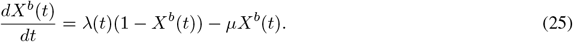

We then separate the variables:

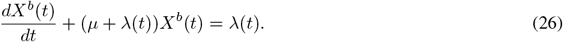

When *λ* is constant over time (i.e.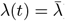), we can solve eq. (26) for the proportion seropositive at time *t* ≥ *b* using the integrating factor approach:

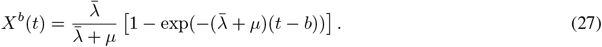

Eq. (27) shows that the proportion seropositive increases monotonically towards a plateau at 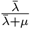, which is below 1(because *µ >* 0).

We can alternatively assume that *λ*(*t*) is piecewise-constant, where each calendar year has a given (constant) FOI. Considering the first year of life for a birth-cohort born at the start of year *B*, we can determine the proportion seropositive by using an integration factor approach:

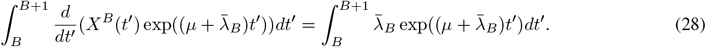

When rearranged, this yields the following expression for the proportion seropositive at the start of year *B* + 1:

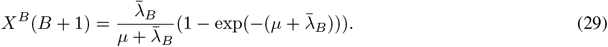

Repeating this exercise for an arbitrary year, *T > B*, produces the following:

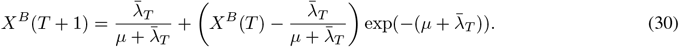

If *µ* = 0, then eq. (30) can be used to produce to eq. (19).

Eq. (30) is an iterative solution, with the proportion seropositive at the end of each year becoming the initial condition for the next piece. The symbolic expression for the solution that results from this iterative process is cumbersome, but it is straightforward to solve for a general *X*^*B*^(*T*), with *T* ≥ *B*, numerically using a for loop (see Algorithm 1).

#### Algorithm 1 R function for solving the time-varying model.

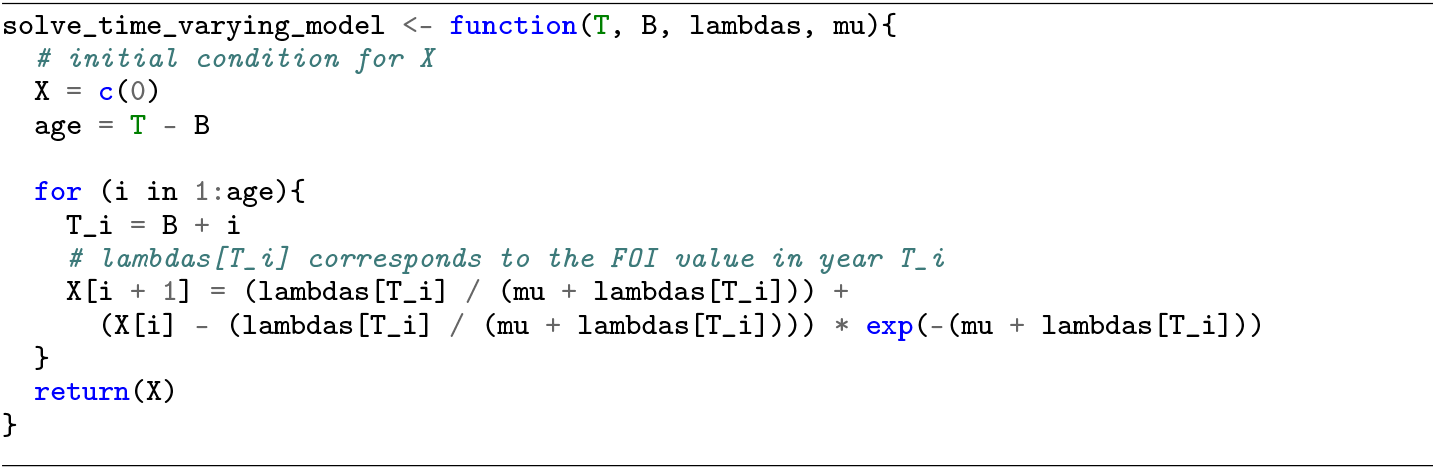

In Figure 4, we show how seropositivity profiles are modified by incorporating seroreversion. Unlike the model without seroreversion, seropositivity of birth-cohorts can decrease over time (Figure 4B): this occurs when the proportion of seropositive individuals becoming seronegative exceeds the proportion of seronegative individuals becoming infected, i.e. if:

**Figure 4:**
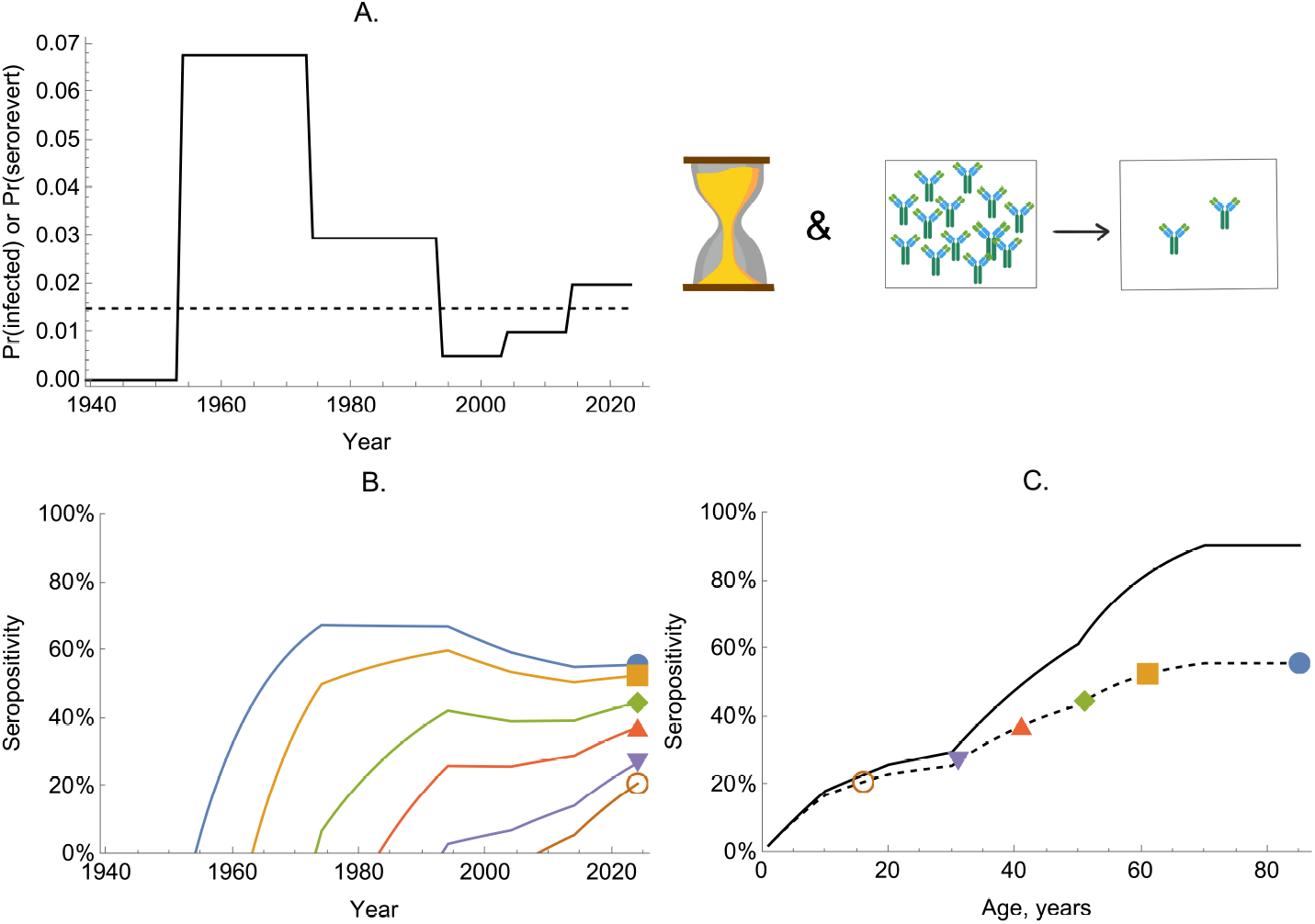
The dynamics of seropositivity in a model with time-varying FOI and seroreversion. Panel A shows the probability of becoming infected per year (given by 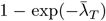) over time (black solid line) and the fixed probability of seroreverting per year (dashed line). Panel B shows the proportion seropositive for six birth-cohorts (coloured lines). Panel C shows the proportion seropositive by age for the population in 2024; the dashed line shows the proportion seropositive for all age groups for the model including seroreversion and the coloured markers correspond to the serpositivities shown in 2024 in panel B; the solid line shows the model solution if the rate of seroreversion were zero.

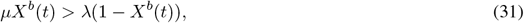

which gives a threshold condition: 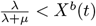.

Overall, the inclusion of seroreversion in the model leads to lower seropositivity at each age (Figure 4C).

In Figure 4B, we see that older cohorts tend to have seropositivity above younger cohorts. We ask, is this generally true for time-varying FOI models with seroreversion? The answer is *nearly*: older cohorts must have seropositivity *at least* as great as younger cohorts. When a new cohort is born, an existing cohort must have seropositivity of zero or higher. From then on, the two cohorts experience the same forces of infection, and differentiating eq. (30) with respect to *X*^*B*^(*T*), we find:

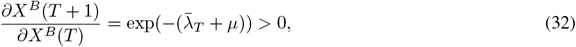

meaning that the cohort starting an interval of fixed 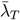 with seropositivity greater than another’s will, throughout the interval, have higher seropositivity. This means that the separate lines representing seropositive fractions through time shown in Figure 4B will never meet.

### 5.2 Real data example: chikungunya virus

Chikungunya virus is widespread in the tropics, where it causes recurrent outbreaks of chikungunya fever. The virus is transmitted by *Aedes aegypti* and *Aedes albopictus* mosquitoes ^9^. Chikungunya fever is characterized by severe arthralgia and myalgia that can persist for years and have considerable detrimental effects on health, quality of life and economic productivity ^9^. Clinical presentation of chikungunya fever is often non-discernible from that of other arboviruses and it can also be confused with other febrile illnesses ^9^. This means that a chikungunya epidemic is likely to go unnoticed or missed, and therefore serosurveys are instrumental in understanding the disease burden or virus epidemiology.

Here, we analyse CHIKV serological data derived from cross-sectional surveys undertaken in 2015 in Burkina Faso and Gabon ^10^ in order to estimate historical transmission patterns. This dataset includes serological information for individuals between 1 and 55 years old (see Figure 5A). Chikungunya is an epidemic disease and to account for this we fit a time-varying serocatalytic model (in a Bayesian framework, see §A.3); our model does not allow for seroreversion.

**Figure 5:**
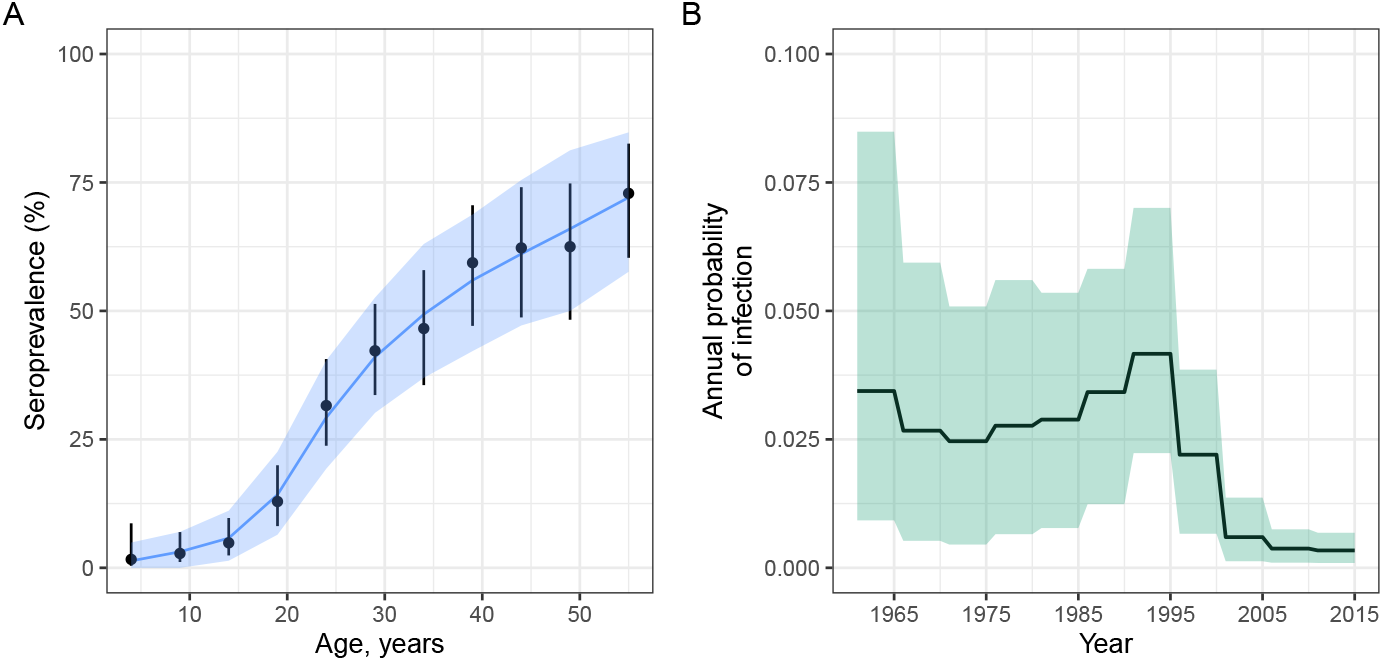
Explaining chikungunya serological data in Burkina Faso and Gabon using a time-varying FOI model. Panel A shows the observed and fitted seroprevalence by age from surveys undertaken in 2015. Points and whiskers represent the observed proportions with 95% confidence intervals. The solid blue line indicates the mean of the posterior samples. Panel B shows the posterior mean annual probability of infection estimates given by 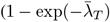. In both panels, the shading indicates the with 95% credible intervals (95% CrIs), representing the 2·5th and 97·5th percentiles of the posterior distributions. We assumed that the FOI was piecewise-constant with pieces of width 5 years.

Serocatalytic modelling Our model provided a reasonable fit to the serological data (Figure 5A). Our reconstructed series for the annual probability of infection (Figure 5B) indicated a likely reduction in transmission more recently, which is similar to that determined in previous work ^10^.

## 6 Age-dependent force of infection

We now explore the case where FOI varies by age but the profile of infection remains constant over time. That is, the FOI experienced by a 5-year-old in 1939 is the same as that experienced by someone of the same age in 2024 but distinct from those aged 10.

### 6.1 Without seroreversion

We first suppose there is no seroreversion meaning the dynamics of seropositivity are described by a slight modification of eq. (4):

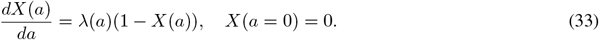

where *a* = *t* − *b* is the age at time *t* of serosurvey for the cohort born at time *b*. We note, however, that eq. (33) uses a different notation to eq. (4), where we no longer use *X*^*b*^; rather, we use *X*, because, if we know the age of individuals, *when* they were born does not influence the trajectories of their seropositivities.

When FOIs are independent of age, the solution becomes:

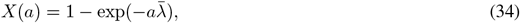

which is the same as eq. (12), because when FOIs do not vary (through time or by age) the time-dependent model and the age-dependent FOI models become equivalent.

Generally, when we assume the FOI varies by year of age (but is constant within each year), the FOI experienced by a cohort of integer age *A* is given by: 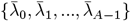.For example, a 3-year-old individual would have experienced three distinct FOIs, each corresponding to one year of their life. In this case, the solution becomes:

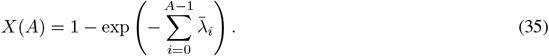

This means that the saturating level of seropositivity is given by: 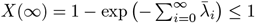.

In Figure 6, we consider a pathogen with transmission patterns similar to those of sexually transmitted infections, which have a strong age-related exposure profile. Here, we model the serological dynamics of such a disease where the FOIs are only dependent on age and not time, i.e., we assume that transmission has been stable through time.

**Figure 6:**
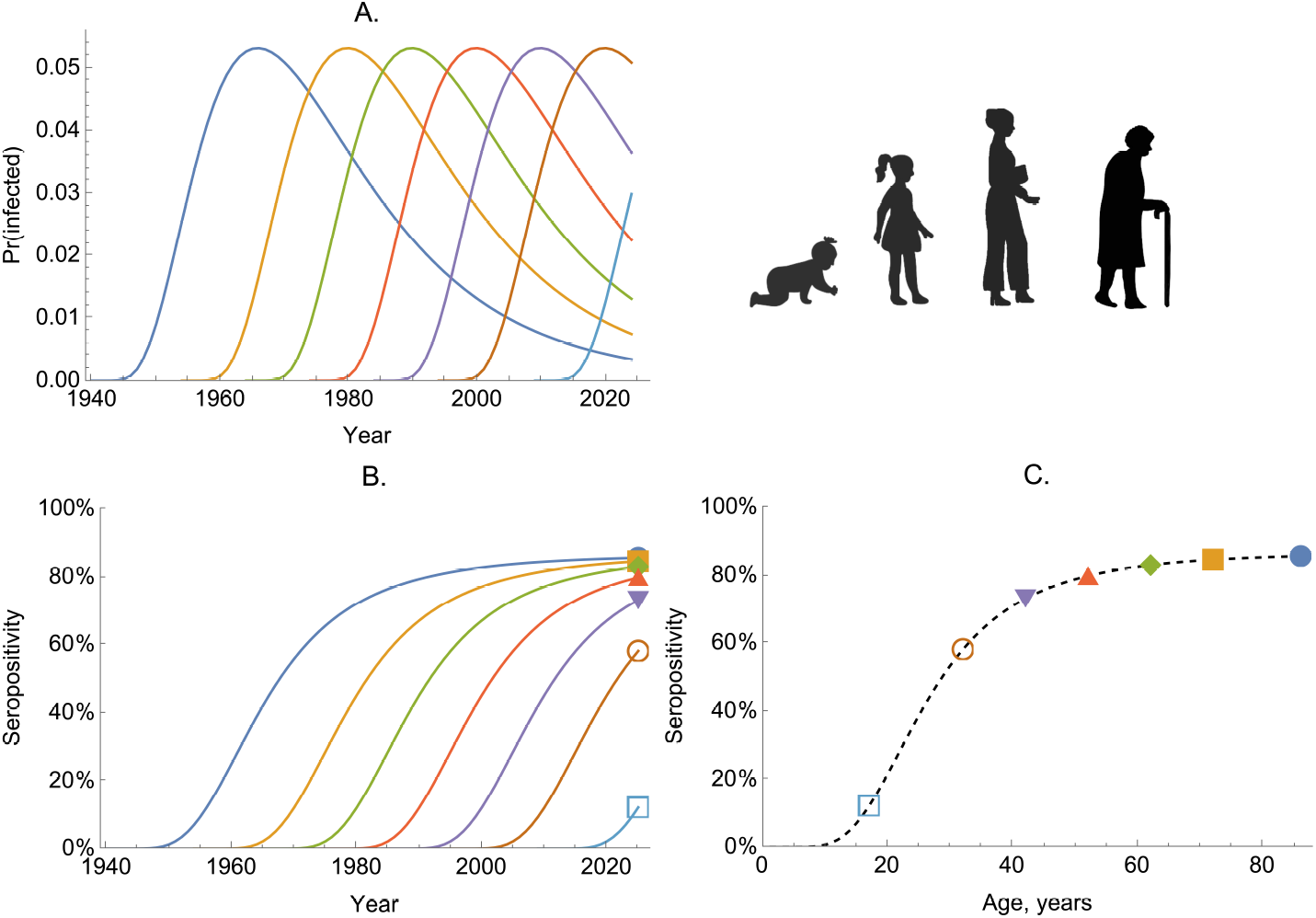
The dynamics of seropositivity with age-dependent infection risk – serological dynamics of a sexually transmitted infection. Panel A shows the probability of becoming infected per year (given by 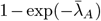for seven birth-cohorts (coloured lines). Panel B shows the proportion seropositive for the same birth-cohorts (coloured lines). Panel C shows the proportion seropositive by age for the population in 2024; the dashed line shows the proportion seropositive for all age groups; the coloured markers correspond to the same 2024 values as shown at the right-edge of panel B.

Specifically, we assume that FOI is highest in those aged in their early 20s and that the age profile of infection risk remains constant over time (Figure 6A). The trajectories of seropositivity follow the same profile, regardless of when a cohort was born (Figure 6B) and older cohorts have higher seropositivity (Figure 6C).

### 6.2 With seroreversion

We now consider an age-dependent FOI model with seroreversion using a slight modification to eq. (33):

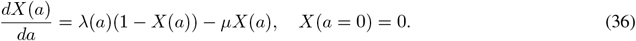

We now solve this equation assuming FOI is piecewise-constant with yearly piece-widths. To do so, we denote integer-valued age by an uppercase *A*:

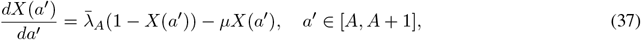

and an integration factor approach can be used to yield:

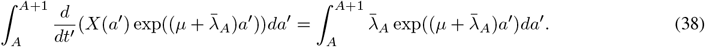

This has the same solution as given by eq. (30) (replacing *T* → *A*):

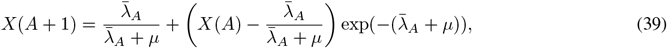

where 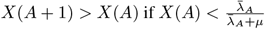.

### 6.3 Real data example: mumps virus

Mumps is a common childhood infection caused by the mumps virus. The hallmark of infection is swelling of the parotid gland, and aseptic meningitis and encephalitis are common complications. Other complications include deafness and pancreatitis ^11^.

We analysed cross-section seroprevalence data collected in 1986-1987 in the UK for individuals between 1 and 44 years of age ^12^ (shown in Figure 7A&C). Like previous work, we assumed that the FOI was age-varying ^12^.

**Figure 7:**
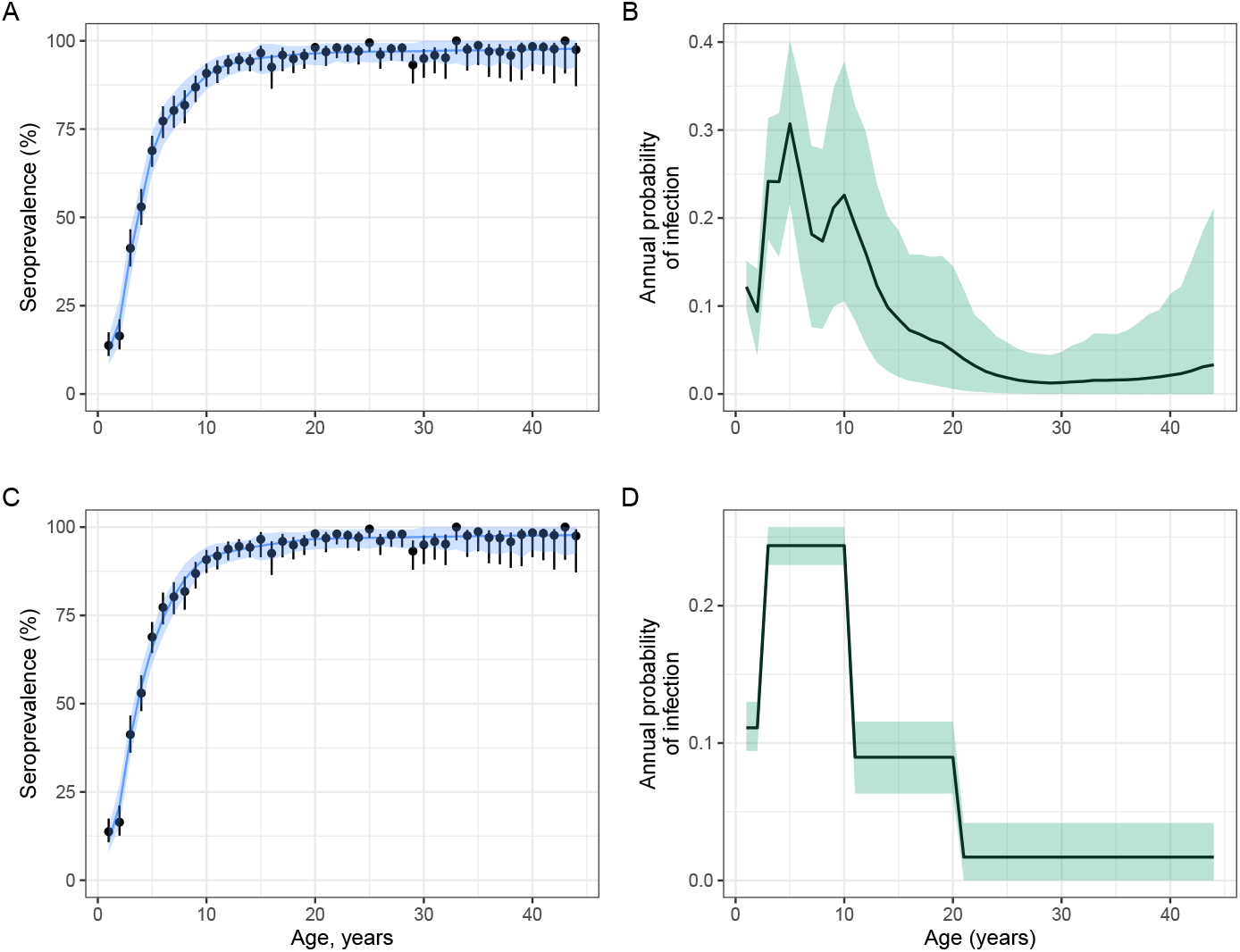
Age-varying FOI model fits for mumps virus. Panels A and C show the observed and model fitted seroprevalence by age. Points and whiskers represent the observed proportions with 95% confidence intervals. The blue lines indicate the mean of posterior samples and the and shading shows the 95% credible intervals representing the 2·5th and 97·5th percentiles of the posterior distributions. Panels B and D show the estimated age-specific annual probability of infection, calculated as 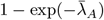,from a model with one-year piecewise-constant FOIs (panel B) and from a model assuming four FOI pieces (panel D). For panels B and D, we estimated four FOI values across the following age bands: (1,2), (3-8), (9,10), (11-44).

We considered two scenarios: (i) piecewise-constant FOIs with one-year age widths, i.e., a distinct FOI for each age class, (ii) four FOI changepoints corresponding to ages 2, 8, 10 and 24 years. The first scenario is a more flexible model assumption but prone to wide uncertainty around each *λ*_*A*_ since there are relatively few data points to inform each FOI. The second scenario characterises transmission intensity across various age-bins – for example, it allows different FOIs in infancy, during pre-school years and during adolescence or teenage years. This scenario may result in lower variance estimates but is at a higher risk of bias.

In both scenarios the model fits were similar and reasonable (Figure 7 A&C). In both models, the inferred FOI peaked in younger children and was low for older age groups. The estimated age-specific annual probability of infection was highest in the age range 2-10 years, with a peak at 0.36 (95% CrI: 0.24-0.52) in scenario (i) (Figure 7B) and 0.25 (95% CrI: 0.26-0.30) in scenario (ii) (Figure 7D). The distinct model fits illustrate the impact of assumptions during inference as shown by higher variance in FOIs in model (i) (Figure 7 C).

## 7. Elevated death rate due to infection

We now suppose individuals exposed to a pathogen have an elevated risk of death. We show that exposure-related mortality biases the observed serological profile and impacts how data should be interpreted. We illustrate this effect by including an additional “deceased (*D*)” compartment to the age-dependent FOI model:

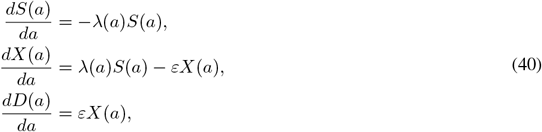

where *ε >* 0 denotes the rate at which previously infected individuals die, and we assume *S*(0) = 1, *X*(0) = 0, *D*(0) = 0 as initial conditions for each birth-cohort. If we consider a special case of eq. (40) for when FOI is time-constant, we can exactly solve the system to give:

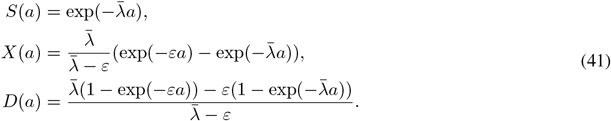

Generally, serological data are collected from survivors. This means that the proportion of those seropositive out of those alive is given by the ratio: 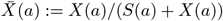.Using eq. (41), we can derive a simplified expression for this quantity as follows:

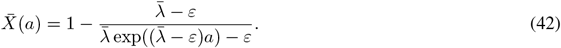

If 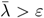,as *a*→ ∞, then 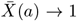;if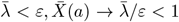. Figure 8 illustrates the analytical solution in eq. (42) in two scenarios: (i) small *ε*, (ii) versus a large *ε*.

**Figure 8:**
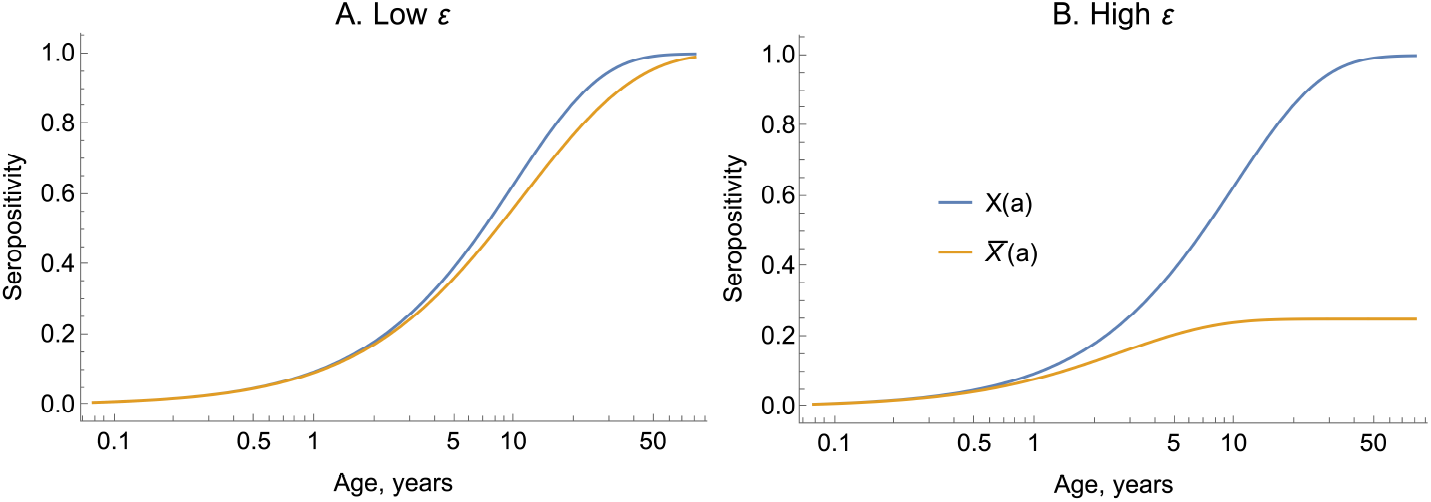
An elevated death rate subsequent to infection can dilute the pool of seropositive individuals. In both panels, we plot seropositivity estimated using eq. (42) (orange lines), which accounts for an elevated death rate post-infection. In panel A, we assume a low death rate, *ε* = 0.05; in panel B, we assume a higher death rate, *ε* = 0.4. In both panels, *λ* = 0.1. The blue lines show the approximate solution given by neglecting to account for the elevated death rate (eq. (13)).

In Figure 8A, we show that when *ε* is small then the exact solution of eq. (42) is nearly equal to the approximate solution which neglects deaths due to infection, 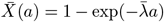.Conversely, when *ε* is large, eq. (42) results in lower 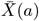 values (orange line, Figure 8B).

### 7.1 Subpopulations with differing mortality risks

An issue with the model described in §7 is that it supposes all those who become infected will die, eventually, due to their infection. However, this is not generally true – usually only a subset of those infected will die due to their infection.

We can represent the heterogeneity in the risk of severe infection leading to death by separating the seropositive populations into two groups: one, *X*_*m*_, comprising individuals experiencing a *mild* infection that does not lead to an elevated death rate; and another, *X*_*s*_, with individuals who experience severe infection and eventually succumb to the infection. The modified model system is then given by:

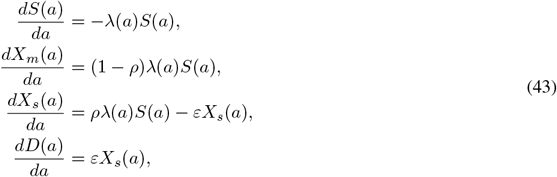

where 0≤ *ρ* ≤1 represents the proportion of infections which are severe (leading to death); alternatively, known as the *infection fatality ratio*.

In this system, the proportion of those living who are seropositive, assuming *λ* is constant, is given by:

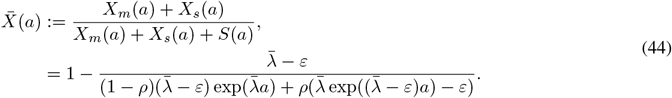

If *ρ* = 1, eq. (44) reduces to eq. (42). If *ρ* = 0, the system becomes equivalent to age-dependent FOI models without death.

Assuming *λ* is piecewise-constant, with one-year pieces, we can determine the proportions susceptible and with mild (m) or severe (s) infection histories at the end of a given piece by:

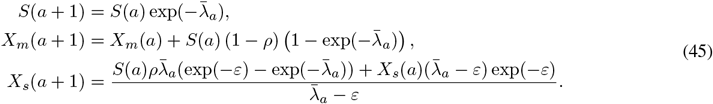

Eqs. (45) can then be used numerically to determine the proportion seropositive of those surviving (i.e., those sampled in a serosurvey). Similarly, eq. (45) is an iterative solution, with the proportion seropositive at the end of each year of life becoming the initial condition for the next piece as shown in Algorithm 2.

#### Algorithm 2 R function for solving the piecewise-constant model described in section 7.1.

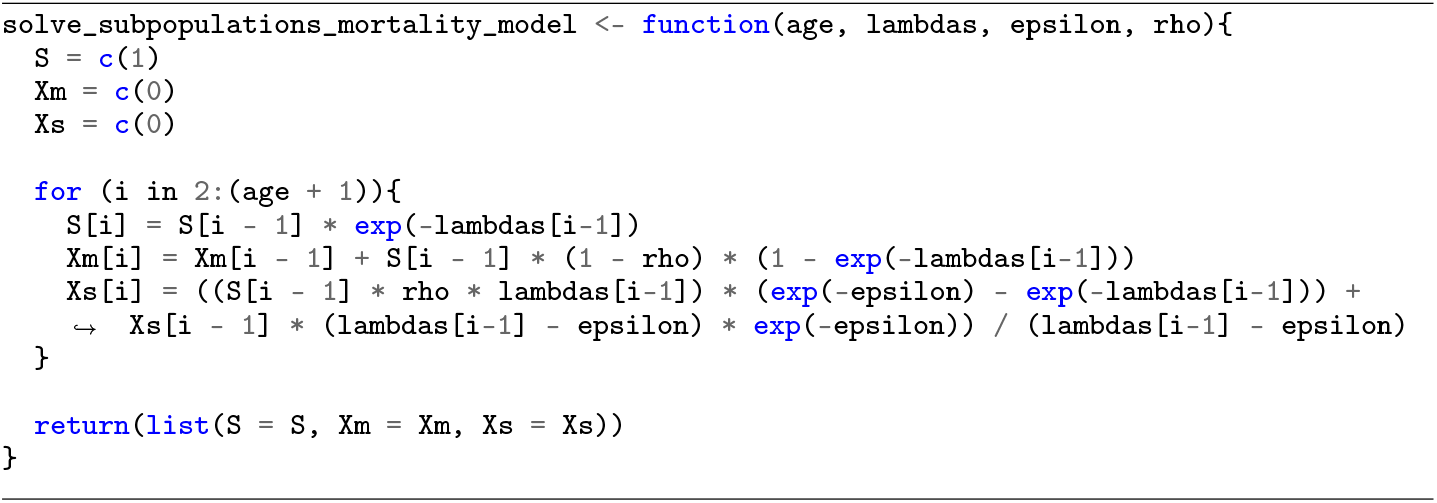

### 7.2 Real data example: Ebola virus

The West African Ebola virus disease outbreak of 2014–16 spread from Guinea to Liberia, Sierra Leone, Mali, Senegal, and Nigeria, and in 28 months, the outbreak had resulted in 28,652 cases and 11,325 deaths ^13^.

We fit the model described in §7.1 to antibody prevalence data from Ebola survivors in Sierra Leone after the 2014-2016 outbreak ^14^. The Sierra Leone study was conducted between 2016 and 2018 and measured IgG to Ebola virus glycoprotein in 1282 individuals of median age 16 years, with an interquartile range of 7–25 years, of whom 107 (8.4%) were seropositive. In this example, we assumed a stepped time-varying FOI, with *λ*=0 before the outbreak in 2014 and an estimated *λ* from 2014 onwards.

For one set of model fits, we set the infection fatality ratio, *ρ* = 0.89^15^ and in another *ρ* = 0. Both models, with and without the assumption of death due to infection, fitted the observed data reasonably (Figure 9A) but estimated substantially different FOIs. The model that incorporated elevated death due to infection estimated an annual probability of infection around six times higher than the model neglecting this (Figure 9B).

**Figure 9:**
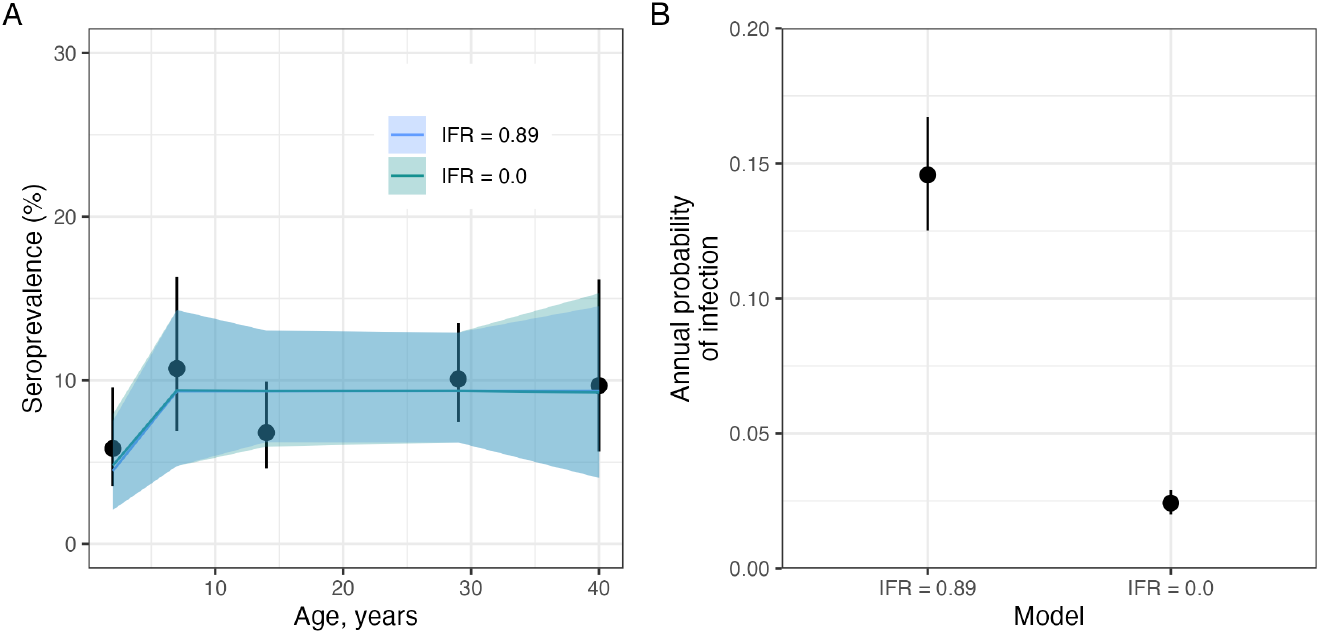
Accounting for an elevated death rate due to infection results in a higher inferred FOI for Ebola virus disease in Sierra Leone during the West African Ebola outbreak in 2014-16. Panel A shows the observed (black circles) seroprevalence with 95% confidence intervals and model-predicted seroprevalence by age (blue and green lines). The predicted seroprevalence (blue and green lines) are derived from two models with and without infection fatality ratio (IFR). Panel B shows the annual probability of infection for the two IFR models.

This example illustrates the importance of accounting for infection-induced mortality in an appreciable fraction of cases – failing to do so can severely underestimate the level of transmission.

## 8 Time- and age-dependent force of infection

Exposure and susceptibility can vary both by age and over calendar time. That is, *λ* = *λ*(*a, t*). Here, we consider a simplified case where the FOI varies as a product of time- and age-specific patterns, *λ*(*a, t*) = *u*(*a*)*v*(*t*), implying persistent age-related patterns that fluctuate in level temporally. A sexually transmitted infection might have FOI following such patterns, where sexual contact rates would typically peak in the early twenties but the transmission rate may be affected by public health policies or treatment and preventative measures, lowering the overall transmission rates across all ages ^16^.

Here, we first consider models without seroreversion. Then the proportion seropositive evolves according to:

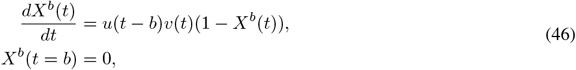

As previously, we assume that *u*(.) and *v*(.) are both piecewise-constant with pieces of width one year; we denote their levels within integer year *T* for a cohort born at the start of year *B* by 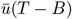 and 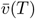.This means that within a year, their product, *λ* is constant, and the proportion seropositive at the end of the year is given by:

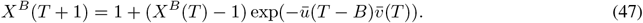

We can then solve for the proportion seropositive at the start of arbitrary year *T > B* for a birth-cohort born at the start of year *B*:

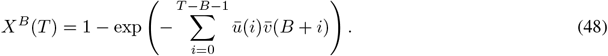

To exemplify the behaviour of this model, we consider a disease with an age-dependent FOI structure similar to that of a sexually transmitted infection: with *u*(.) peaking in the early 20s. At the same time, we allow a rapid increase in the level of transmission, starting in the year 1980 (Figure 10A), for example, representing a widespread increase in the number of infected individuals. This upward shift in transmission means that the individuals with peak sexual activity during the period of elevated transmission experience a disproportionately higher risk of infection and the seropositive proportion of these cohorts can exceed those of older individuals (Figure 10 B&C).

**Figure 10:**
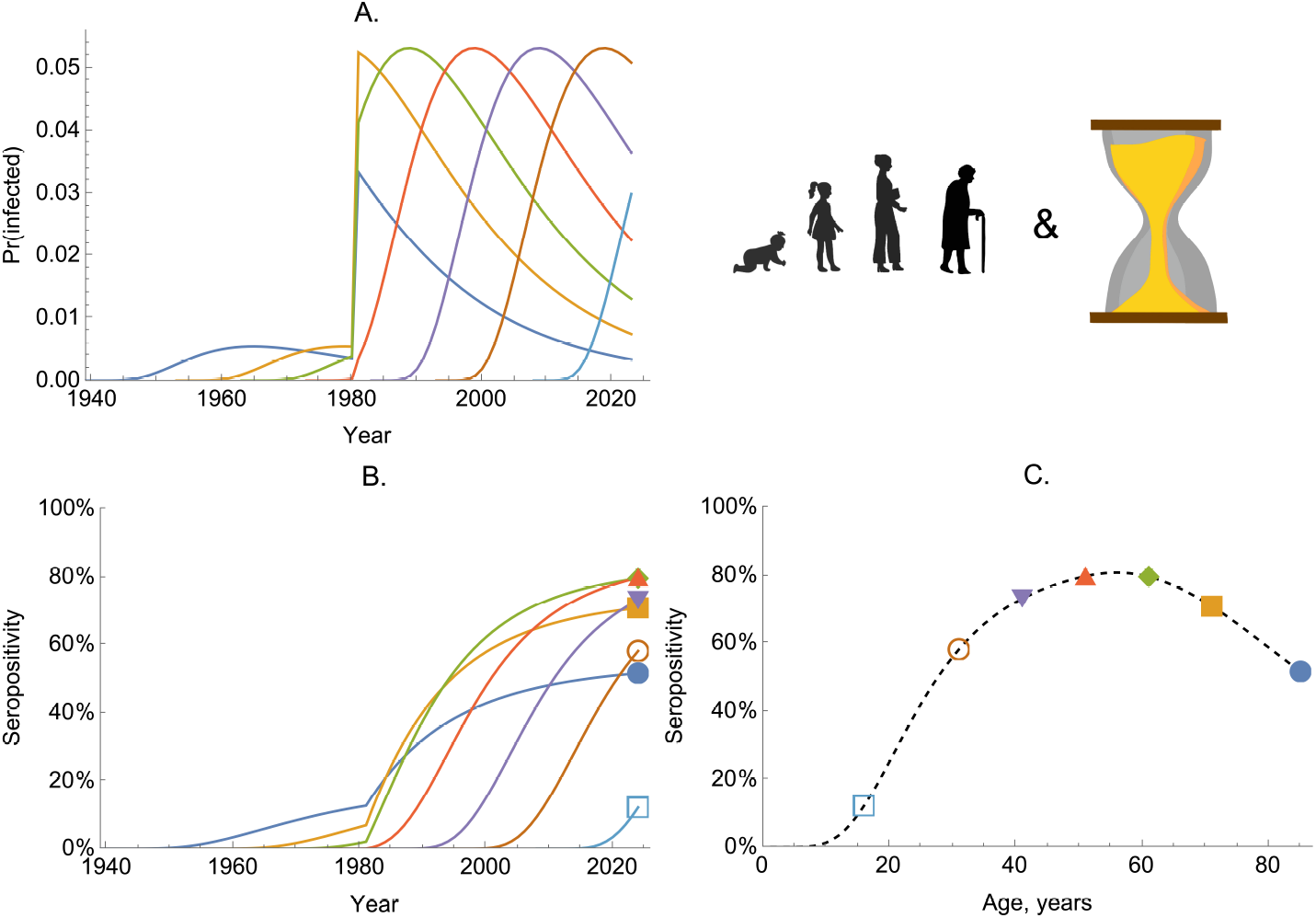
The dynamics of seropositivity with age- and time-dependent infection risk. Panel A shows the probability of becoming infected per year (given by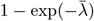) for seven birth-cohorts (coloured lines). Panel B shows the proportion seropositive for the same birth-cohorts (coloured lines). Panel C shows the proportion seropositive by age in a single cross-section, taken in 2024. The dashed line shows the proportion seropositive across all age groups, while the coloured markers correspond to the same values shown in 2024 in panel B.

### 8.1 Including seroreversion

If we allow seroreversion, the seropositivity at start of year *T* is given by (through analogy to eq. (30)):

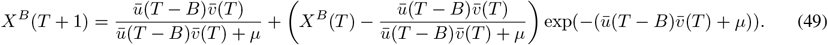

Like for the time- and age-only models, when *u*(.) and *v*(.) vary by age and year, there is no simple analytical expression for the seropositivity. Nonetheless, eq. (49) can be used to produce an iterative solution if we assume *u*(.) and *v*(.) are piecewise-constant.

### 8.2 Real data example: HIV

The force of infection for HIV likely varies over time and by age. Additionally, the majority of infected individuals bar a few rare cases remain infected throughout their lives. As a result, we can model HIV infection prevalence in the same way we model seropositivity for other pathogens, using models without seroreversion.

In the absence of treatment, HIV infection eventually leads to death, and so it is crucial to account for infection-related mortality when analysing HIV prevalence data. Additionally, it is important to account for the delay from infection to death. Progression to AIDS and death typically follows a long asymptomatic period – estimated to be around 10 years ^17^. To model HIV serodynamics, we use the following system:

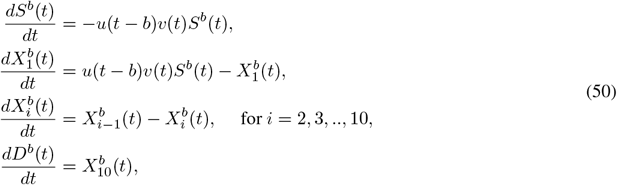

where *S*^*b*^(*b*) = 1 and all other compartments have initial conditions set to zero. In Eqs. (50), we have made use of the linear-chain trick (see, for example, ^6^) by splitting the HIV infection-positive individuals into 10 compartments: *X*_1_, …, *X*_10_, and death occurs only after the last compartment is reached. Because the rate parameters in front of each *X*^*b*^ term are 1 in eq. (50), this results in a typical duration to death due to infection following a gamma(10, 1) distribution, which has a mean of 10 years (see ^3^).

As before, we assume that *u*(.) and *v*(.) are piecewise-constant with one-year pieces. Since the system is linear, we can write it as a vector differential equation by defining 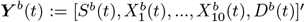 as a vector of states and 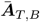 as a matrix of constants that are fixed for a given calendar year *T* assuming individuals were born at the start of year *B*:

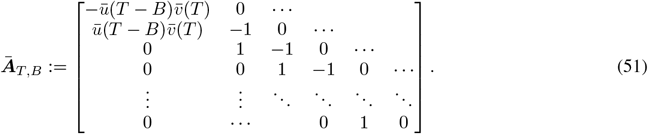

Using this matrix, we can write down the system of equations as a single vector equation:

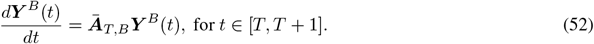

We can then analytically solve eq. (52) to yield the seroprevalence at the start of year *T* + 1 of those born at the start of year *B* (note that *T* and *B* are integers):

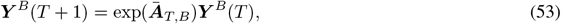

where, here, exp(.) denotes the matrix exponential. The matrix form means that a general solution for the system can be written down as follows:

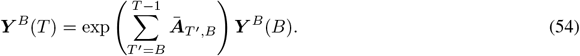

Since all serocatalytic models described here are linear ordinary differential equation systems, we can solve such models this way; however, this abstract form generally does not permit such mathematical analysis as the more bespoke methods presented up to this point.

We fit the model structure described by Eqs. (50) to age-specific HIV prevalence data from a HIV survey conducted in rural KwaZulu-Natal, South Africa, in 2003^18^ (points and uncertainty bars in Figure 11A). The survey was conducted just before large-scale roll-out of HIV care and antiretroviral treatment (ART) in 2004 and before substantial reduction of HIV-associated mortality and increased life expectancy ^18^. The data include HIV-seroprevalence profiles of women aged 15–49 years. Older age groups did not commonly participate in the HIV surveillance and were only routinely included from 2007 onwards ^18^.

**Figure 11:**
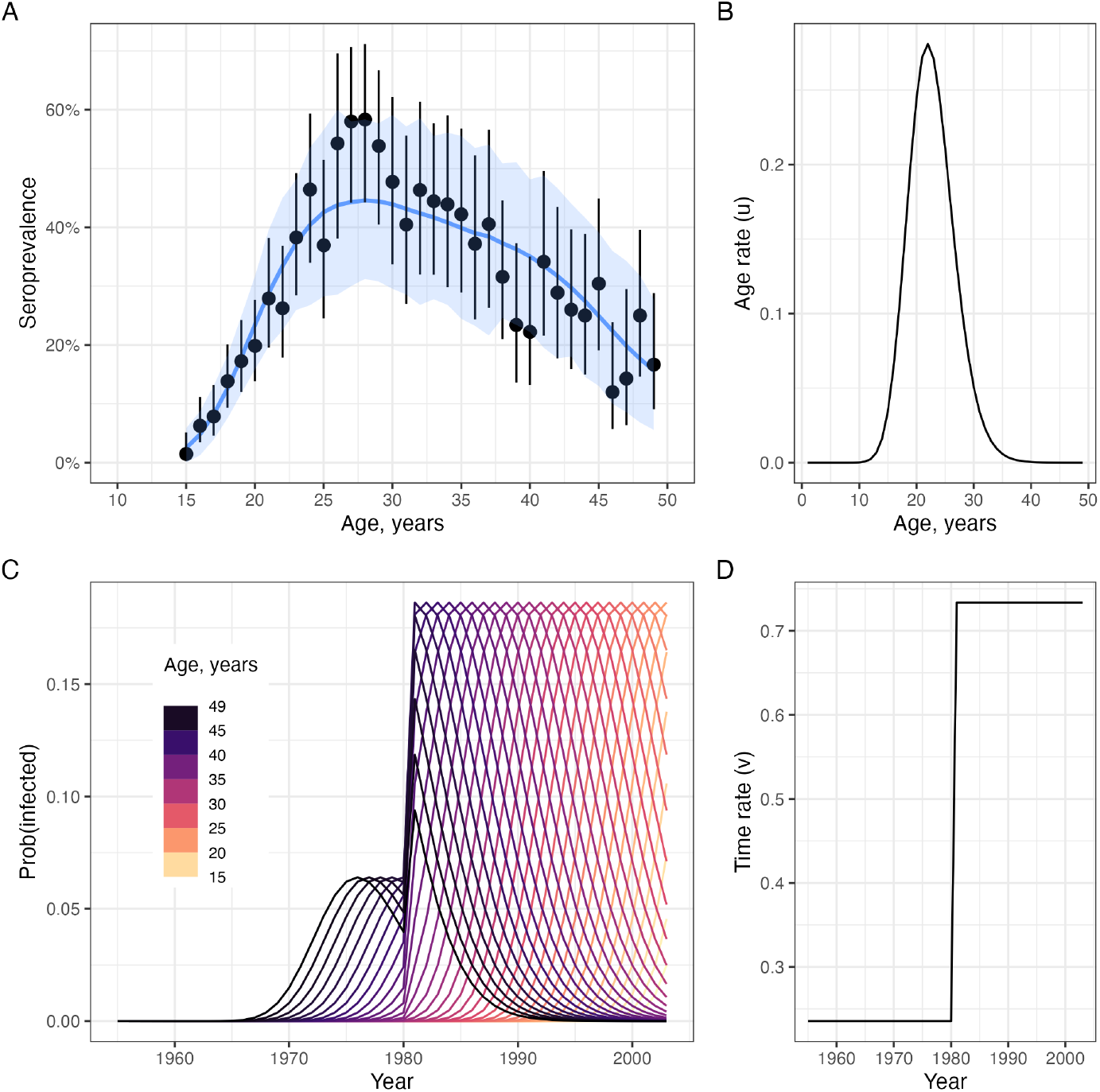
Modelling HIV prevalence in South Africa before widespread treatment. Panel A shows the observed and model fitted seroprevalence by age. Points and whiskers represent the observed prevalence with exact 95% confidence intervals. The blue line and shading indicate the mean of posterior samples, with 95% credible intervals representing the 2·5th and 97·5th percentiles of the model’s posterior distributions. Panel B shows the estimated age patterns of infection *u*(.). Panel C shows the annual probability of becoming infected for each cohort. The probability of infection is calculated as 1 − exp(−(*u*(.)*v*(.))). (D) shows the estimated time patterns of infection *v*(.).

The modelled seroprevalence estimates are in broad agreement and overlap with the observed data (Figure 11A).

The age of peak infection risk was estimated to lie between 20 and 30 years (Figure 11B); this is consistent with patterns of sexual behaviour and partnership observed in longitudinal population-based surveys ^19^.

We simplified the assumptions about changes in disease transmission over time by allowing one transmission rate multiplier prior to the year 1980 (*v*(*T*≤ 1980)) and another after 1980 (*v*(*T >* 1980)). Our model estimates indicated a roughly 3-fold increase in transmission levels between these two periods (Figure 11C× D), from *v*(*T* ≤ 1980) = 0.23 to *v*(*T >* 1980) = 0.73, resulting in a commensurate increase in the probability of infection by age 40.

## 9 Maternal antibodies

For many pathogens, offsprings inherit antibodies from their mothers. So-called maternal antibodies are typically short-lived, yet failing to account for their influence in serological measurements can bias FOI estimates, as we now illustrate.

We now consider a system where FOIs are age-dependent only and with no seroreversion, and we illustrate how the impact maternal antibodies can be modelled. To do so, we assume that only after individuals have cleared their maternal antibodies can they become naturally infected, and that the rate of loss of maternal antibodies is independent of age. This results in a modified system of equations:

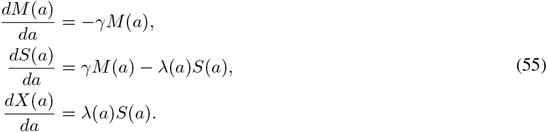

where *M* (*a*) is the proportion of the population with maternal antibodies of age *a*.

For initial conditions, we assume that *M* (0) = 1, *S*(0) = 0, *X*(0) = 0. Assuming that all births have maternal antibodies for a particular pathogen may be crude, and we discuss this in §9.2.

We first consider an idealised scenario when the FOI is constant. Then the above system has the following solution:

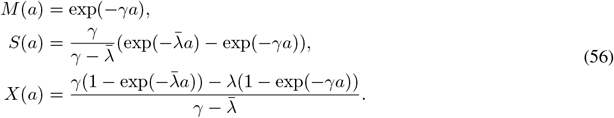

The rate of removal of maternal antibodies is typically high and will initially dominate the dynamics. The FOI will generally be much lower than this rate (i.e.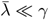) meaning 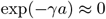 for older age cohorts, and the proportion seropositive will follow:

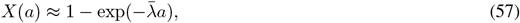

which reproduces eq. (34).

For many pathogens, the typical duration of maternal antibodies is around 6 months, meaning *γ* = 2 per year. In Figure 12, we consider two scenarios corresponding to a 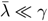 (panel A) and 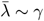 (panel B), where we assume *γ* = 2. We also plot the approximate solution given by eq. (57), which neglects the influence of maternal antibodies. This shows that when 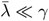.it makes little difference to the modelled seropositive proportion to neglect the effect of maternal antibodies, but when 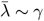, this induces bias in this proportion.

**Figure 12:**
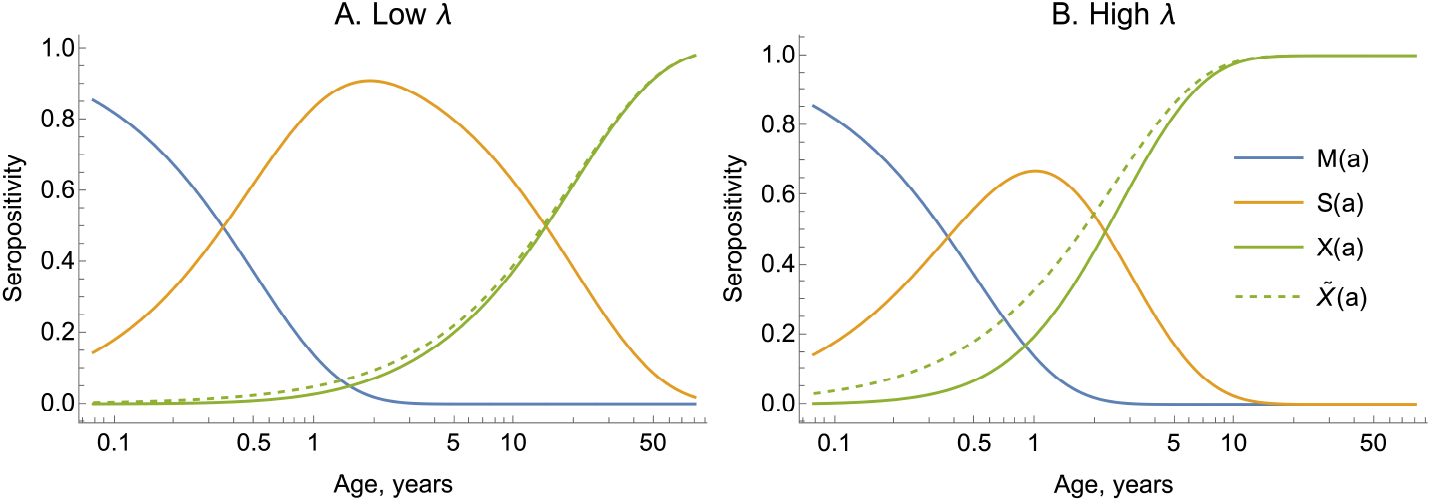
Neglecting maternal antibody dynamics can bias the proportion seropositive. In both panels, we plot Eqs. (56) (solid lines), where we assume *γ* = 2 per year; in panel A, we assume *λ* = 0.05; in panel B, we assume *λ* = 0.4. The dashed lines shows the approximate solution given by neglecting the effect of maternal antibodies (i.e. eq. (57)). Note, the horizontal axis is on the log scale, which cannot start at zero and the reason the maternal compartment does not begin at *M* (0) = 1.

We typically cannot differentiate the actual proportion with maternal immunity from those with immunity due to infection during their lifetimes, which means that we can consider the overall proportion seropositive as:

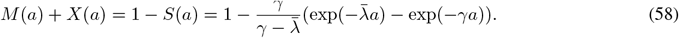

### 9.1 Real data example: enterovirus D68

Enterovirus D68 (EV-D68) infection leads to severe acute respiratory distress in children below 5 years of age with clinical symptoms of hypoxia and wheezing associated with a significant increase in pediatric hospitalisations ^20^. A subset of children develop central nervous system complications and in serious cases respiratory failure ^20^. We used EV-D68 serological data to illustrate the effect of neglecting maternal antibodies in estimation of seroprevalence and the FOI. The data consisted of individuals with ages ranging from newborns to 80 years old, sampled in the UK in 2006^21^. Although recent studies have suggested there may have been increased EV-D68 transmission over time, particularly after the EV-D68 emergence in 2014^22,23^, there had not been a major EV-D68 outbreak reported in the UK at or before 2006. So, we assumed age- and time-independent risk of infection for the population sampled.

We compared two models: (i) an age- and time-independent model that considers the presence of maternal antibodies – as in eq. (56), (ii) and one that does not – eq. (57). The model in eq. (56) fitted the observed seroprevalence data well for lower age groups (ages 1-5 years) compared to the model defined in eq. (57) (Figure 13). We estimated (i) *γ* = 1.6 per year (95% CrI: 1.0-2.9) suggesting an average duration of maternal antibodies of 234 days or around 8 months, and (ii) an annual probability of infection of 0.18 (95% CrI: 0.15-0.20).

**Figure 13:**
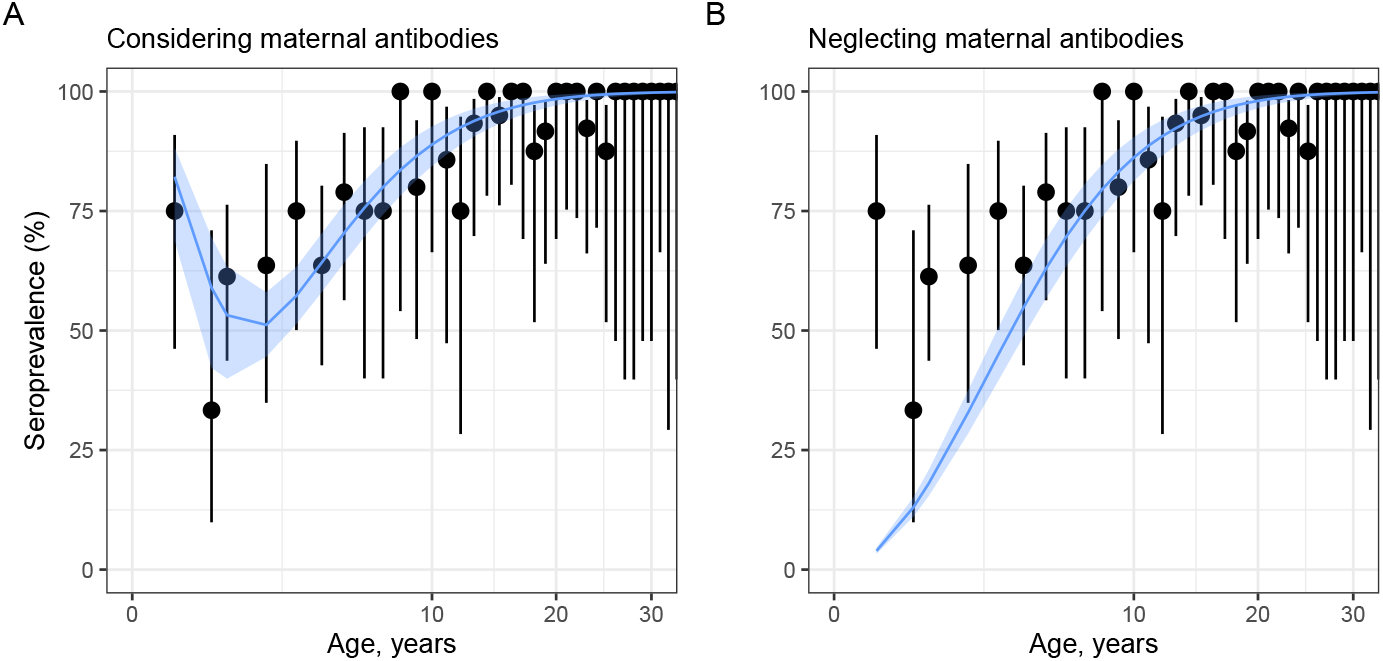
Accounting for maternal antibody dynamics affects the inferred seroprevalence for enterovirus D68. Both panels show the observed (black points, with 2.5th and 97.5th percentiles shown as vertical error bars) and model-predicted (blue lines) seroprevalence by age. The 95% credible intervals (95% CrIs) representing the 2.5th and 97.5th percentiles of the model’s posterior distributions are shown as the blue shading. Panel A shows the results from a model considering maternal antibody dynamics, and panel B shows the model fit when neglecting maternal antibody dynamics.

### 9.2 Feedback between mothers and their babies

Whether a child has maternal immunity depends on the mother’s history of exposure to a pathogen and if the mother retained transferable antibodies. This in turn depends on maternal age and the birth year of the mother. These can be embodied in the initial condition of the system:

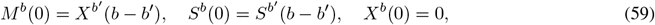

where *b*^′^ *< b* is the date/time when the mother was born meaning *b* − *b*^′^ *>* 0 is the age at which they give birth. Accounting for maternal age at birth might be most relevant if there are substantial changes in seroprevalence around the child-bearing age as well as considerable FOI in the early years of life.

## 10 Discussion

This paper illustrates uses of serocatalytic models to form epidemiological insights about pathogen transmission patterns. Whilst our paper surveyed a range of serocatalytic models, there are a host of other developments of these models that, for brevity, we omitted. This includes how to account for vaccination (e.g., ^24^, ^25^), the impact of migration and immigration (e.g., ^26^, ^27^, ^28^), seasonality (^29^), cross-strain immunity (^30^, ^31^) and cross-reactivity of immunological assays (e.g., ^32^). We also did not discuss how the basic reproduction value can be inferred by fitting serocatalytic models ^33^. We aim to cover these topics in a follow-up article.

For epidemiological studies, these models are typically fitted to serological data, and there are many nuances for doing so. We have developed an open-source R package, serofoi ^4^, a fully tested software package for fitting serocatalytic models to data while incorporating best practices for model inference. A recurring theme when considering inference for these models is that the conclusions drawn are highly contingent on the assumptions made. This is because often divergent theories of disease circulation can generate similar seroprevalence patterns. Epidemiological context is then crucial to ensure that serological data are correctly interpreted.

## Data Availability

All data analyzed are available at https://github.com/ekamau/serocatalytic_models.

https://arianajunjie.github.io/seropackage/

https://github.com/ekamau/serocatalytic_models

## 11 Notes

### 11.1 Ethics approval

Not applicable

## 11.3 Acknowledgements

SB would also like to thank Merton College, University of Oxford where she is the Peter J Braam Early Career Research Fellow in Global Wellbeing.

## A.1 Muench’s serosurvey data for yellow fever and methods for their analysis

Muench’s serological datasets for yellow fever are contained within ^2^: for the two Brazil datasets, in their Chart 1; and for the Colombia dataset, in their Table 1. The data from the charts were digitised using *webplotdigitizer* ^34^. The chart-derived data had no information on the sample sizes, unlike the tabular data.

For all datasets, we used the method of maximum likelihood to estimate the FOIs. We used two different serocatalytic models to analyse the data and estimate the FOI.

### Brazil datasets

We assumed an age- and time-constant FOI, 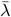,meaning the proportion of individuals seropositive by integer age *A* was given by:

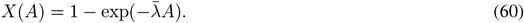

Assuming counts of *x*(*A*) seropositive individuals from a sample of *n*(*A*) individuals of integer age *A*, this implies the probability of observing those data given by:

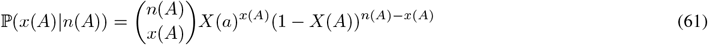

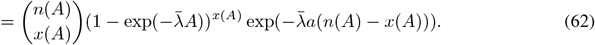

Considering a sample of *n* distinct age categories, where in age-category *i* (with individuals of age *a*_*i*_) we observe *x*_*i*_ seropositive individuals out of *n*_*i*_ individuals tested, we can write down the likelihood as:

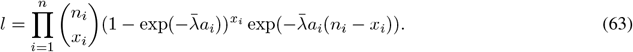

From eq. (63), we can write down the log-likelihood up to a constant:

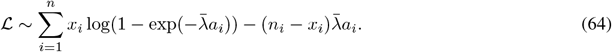

Taking the derivative of eq. (64) and setting this to zero, we have the condition satisfied by the maximum likelihood estimator:

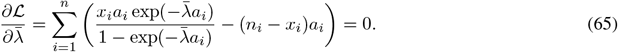

**Glossary S1:**
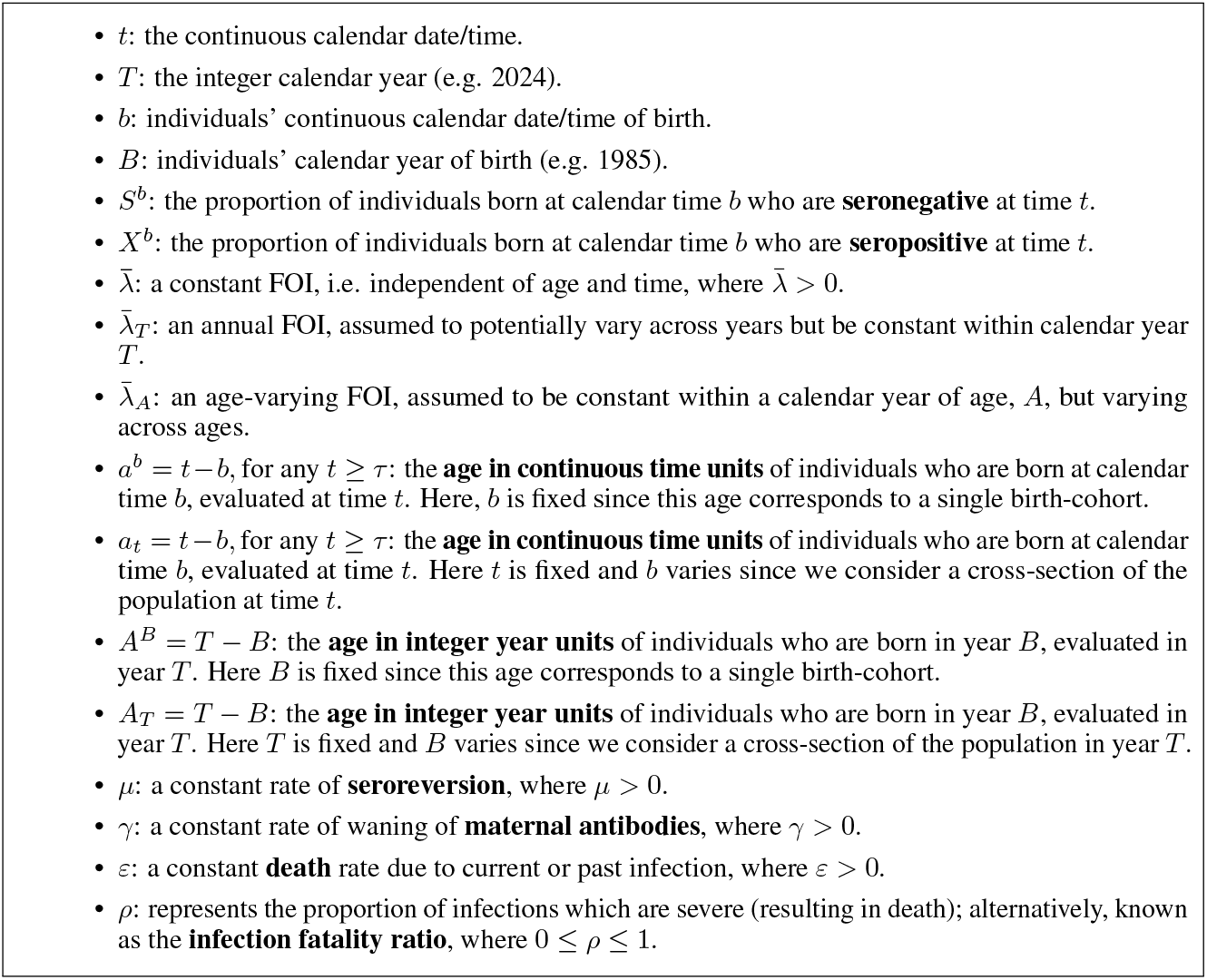
Mathematical notations used in this paper.

Eq. (65) contains *n*_*i*_, which is unknown for the Brazil datasets. But the Brazil data contained only the proportions seropositive, *p*_*i*_, and we assume that *x*_*i*_ = *p*_*i*_*n*_*i*_; substituting this into eq. (65) leaves a factor *n*_*i*_ in all terms which can be divided through by.

The solution to eq. (65) can only be determined numerically, and we used R’s *optim* function to locate the maximum likelihood estimates for each of the two datasets.

### Colombia dataset

We assumed that anyone alive at the time of the 1929 yellow fever epidemic was equally likely to be exposed to the virus and experienced an FOI, *λ*. We assume the serosurvey was conducted *τ* years after the epidemic. This results in a proportion seropositive given by:

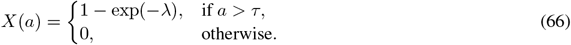

The youngest age group in the dataset is 5-9 and since the study was published in 1934, we assume that everyone could have been exposed to the virus (that is *a > τ* for everyone in the sample).

Through similar logic to before, we can determine a condition for the maximum likelihood estimator, 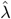,although this time in closed form:

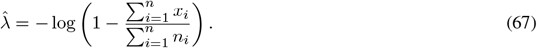

## A.2 Transmission dynamics model including waning immunity

To generate Figure 2, we simulated from a transmission dynamics model of the following form:

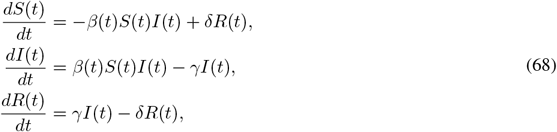

where *δ* = 0.002 per year is the rate at which recovered individuals become susceptible again (i.e. their immunity wanes), and *γ* = 0.05 per year is the rate at which infected individuals recover. We assume that the rate of infection, *β*(*t*), is piecewise-constant and given by:

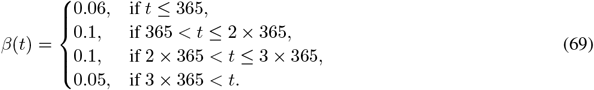

The initial conditions for the system given by eqs. 68 were: *S*(0) = 0.99, *I*(0) = 0.01, *R*(0) = 0, and the system was solved numerically using Mathematica’s inbuilt NDSolve method.

### A.3 Bayesian inference for serocatalytic models

Throughout this paper, we show that serocatalytic models can be fitted to age-structured seroprevalence data to yield estimates of FOIs and other quantities.

Here, we provide an overview of the approach used for inference in this paper. In each of the real data examples, our dataset consisted of serosurveys conducted at a specific time (which we assume is at the start of integer year *T*). The serosurvey results can be summarised by a series of pairs (*x*_*A*_, *n*_*A*_), where *x*_*A*_ denotes the count of seropositive individuals of integer age *A* within those sampled, *n*_*A*_. Throughout, we assume that the proportion seropositive follows:

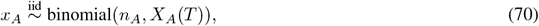

where *X*_*A*_(*T*) is the solution to a serocatalytic model and represents the seropositivity for individuals of age *A* at the start of calendar year *T* in the population where the survey was conducted.

In some of the analyses, we assumed that the FOIs are piecewise-constant; in others, we assumed that the FOI followed a parametric curve and inferred the parameters which specified that curve. In Table S1, we describe the assumptions made for each of the model fits.

We used a Bayesian framework for inference, which requires prior distributions to be specified for all unknown model parameters. In Table S1, we provide the prior distributions used in each of the analyses: these were chosen to allow a wide range of model solutions.

To fit the models, we used Markov chain Monte Carlo (MCMC) sampling through Stan’s default NUTS algorithm ^35^. For each model fit, we used ≥ 3000 MCMC iterations per chain across four Markov chains. Model convergence was diagnosed by 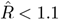 and effective sample sizes (bulk and talk) above 200 for all parameters.

**Table S1:** Priors used in model fitting. The right column lists the respective section in the paper where the model is fitted to a real serosurvey dataset. The parameter *ε* in the *elevated death rate due to infection* model is given by *ε* = 1*/κ*, where *κ >* 0 represents the typical time until death. The age-component of the FOI in the *time- and age-dependent* model is parametrically modelled as a function of *a, b* and *c* parameters shown in this table: *λ* = *c* ×gamma(*age*| *a, b*), where gamma(*age* | *a, b*) is a gamma distribution probability density function evaluated at age, which has mean *a/b*; the time component in the FOI is given a uniform prior between 0 and 1. The *λ*_*t*_ in section §5.2 and *λ*_*a*_ in section §6.3 (panels B and D in Figure 7) were divided into smaller groups, or “chunks”, and the FOI for each chunk was estimated. See https://github.com/ekamau/serocatalytic_models for further details.

| Model | Priors | Corresponding section |
| --- | --- | --- |
| Time-varying FOI, $\lambda_i$,<br>where $i$ is the index of an FOI piece | $\log(\lambda_{i>1}) \sim \text{Student-t}(\nu, \log(\lambda_{i-1}), \sigma)$,<br>$\log(\lambda_{i=1}) \sim \text{normal}(-3, 1)$,<br>$\sigma \sim \text{Cauchy}^+(0, 1)$,<br>$\nu \sim \text{Cauchy}^+(0, 1)$. | §5.2 |
| Age-varying FOI, $\lambda_i$,<br>where $i$ is the index of an FOI piece | $\log(\lambda_{i>1}) \sim \text{normal}(\log(\lambda_{i-1}), \sigma)$,<br>$\log(\lambda_{i=1}) \sim \text{normal}(-3, 1)$,<br>$\sigma \sim \text{Cauchy}^+(0, 1)$. | §6.3 |
| Elevated death rate due to infection | $\lambda \sim \text{exponential}(1)$,<br>$\kappa \sim \text{normal}^+(0.04, 0.02)$. | §7.2 |
| Time- and age-dependent FOI | $a \sim \text{Cauchy}^+(0, 1)$,<br>$b \sim \text{Cauchy}^+(0, 1)$,<br>$c \sim \text{Cauchy}^+(0, 1)$ | §8.2 |
| Maternal antibodies | $\lambda \sim \text{exponential}(1)$,<br>$\gamma \sim \text{Cauchy}^+(0, 1)$. | §9.1 |

### Footnotes

1 This neglects the effect of maternal antibodies, which we discuss in §6.

2 In the stochastic individual-based version of this model.

3 https://github.com/epiverse-trace/serofoi

